# DiabetIA: A Longitudinal Electronic Health Record Dataset for Diabetes Research from Mexico

**DOI:** 10.64898/2026.11.24.26368871

**Authors:** Arturo López-Pineda, Joaquin Tripp, Daniel Santana-Quinteros, Marisol Flores-Garrido, Luis M. García-Velázquez, Karina M. Figueroa-Mora, Anel Gómez-García, Cleto Alvarez-Aguilar

**Author notes:** Corresponding author Address correspondence to: **Dr. Anel Gomez-García**, Associate Researcher D, Biomedical Research Center of Michoacan (CIBIMI), Decentralized Administrative Operation Office (OOAD) Michoacan Mexican Social Security Institute (IMSS), **Dr. Cleto Alvarez-Aguilar**, Associate Professor, Faculty of Medical and Biological Sciences “Dr. Ignacio Chávez”. Universidad Michoacana de San Nicolás de Hidalgo (UMSNH), Morelia, Michoacan, Mexico.

## Abstract

Longitudinal electronic health records (EHRs) from primary care are scarce in low- and middle-income countries, and few have been made available to researchers in Latin America. We present DiabetIA, a longitudinal dataset derived from the primary care records of the Mexican Institute of Social Security (IMSS). The first release comprises 136,674 adults followed in six IMSS family medicine units in the state of Michoacan between 2005 and 2020, with more than 3.2 million visits summarized in 414,770 two-year windows with at least one visit. Each window includes demographics, ICD-10 diagnoses, anthropometry, vital signs, laboratory results and prescriptions, and type 2 diabetes diagnoses in the following year as prediction targets. At baseline, 24.4% of the patients had type 2 diabetes, 37.1% hypertension and 32.3% obesity. Technical validation assessed completeness, plausibility, consistency, temporal coverage and agreement with a state health survey. DiabetIA is freely available for download to support research on chronic disease trajectories and prediction in an underrepresented population.

## Background & Summary

Electronic health records (EHRs) have emerged as one of the most powerful resources for biomedical research in the real world. When systematically curated, de-identified, and shared with the research community, EHRs enable a longitudinal perspective on health and disease, capturing not only diagnoses and treatments but the full arc of patient care across time^1^. Unlike clinical trials, which are limited by selective patient eligibility and statistical constraints defined by the trial designers, EHR data reflects the complex, continuous nature of healthcare delivery, spanning preventive care, chronic disease management, and acute interventions over months, years, or even decades^2^. Primary care records are a particularly valuable source of real-world data (RWD), because they follow the same patients for many years and capture the prevention and management of chronic diseases in the general population^3^.

Creating longitudinal EHR datasets ready for global biomedical research poses substantial challenges^3^. Clinical records are often siloed, scattered across paper and electronic trails, and often with free-text and files lacking structured data. Transforming this raw information into high-quality, de-identified, and interoperable formats requires technical expertise, rigorous quality control, and institutional collaboration. As a result, RWD of this kind, particularly from healthcare systems in middle-income countries, is rarely shared openly. Yet when made accessible, RWD can be transformed into real-world evidence (RWE), offering insights into disease burden, treatment effectiveness, healthcare disparities, and system performance under routine conditions^4^.

Clinical datasets such as the Medical Information Mart for Intensive Care (MIMIC-IV)^5^ and the eICU Collaborative Research Database^6^, and platforms such as OpenSAFELY^7^ in England, have demonstrated the scientific value of structured EHRs, particularly in hospital-based and critical care environments. These resources have enabled major advances in clinical epidemiology, machine learning, and reproducibility. However, they are largely limited to acute-care settings and short-term encounters, offering only a narrow view of the broader healthcare continuum. In contrast, primary care represents the first and most sustained point of contact between patients and any health system, playing a central role in prevention, chronic disease management, and population health^8^. Despite its importance, primary care data remains underrepresented among the EHR datasets available to researchers.

At the same time, large-scale biobanks such as the UK Biobank^9^, All of Us^10^, or FinnGen^11^ have made landmark contributions to genomics and population health research by linking biospecimens with survey-based phenotypes and limited clinical summaries. These initiatives have supported genome-wide association studies and polygenic risk modeling at scale, yet they rarely include comprehensive, longitudinal clinical records derived from routine care^12^. Similarly, disease-specific registries such as the Surveillance, Epidemiology, and End Results (SEER) Program^13^ provide valuable cancer incidence data but lack real-time treatment records, comorbidities, or non-oncologic health information. For researchers seeking to study real-world disease progression, treatment pathways, and health system utilization, there is a critical need for encounter-level, longitudinal clinical data that captures patient trajectories in primary care settings.

Latin America remains one of the most underrepresented regions in the global landscape of clinical data available for research, despite being home to large national health systems, such as Brazil’s Sistema Único de Saúde (SUS), which serves over 160 million people^14^, and Mexico’s Instituto Mexicano del Seguro Social (IMSS), which covers more than 70 million^15^. This absence is particularly notable given the region’s persistent and disproportionate burden of non-communicable diseases. Chronic conditions such as obesity, type 2 diabetes, hypertension, and other cardiovascular diseases are among the leading causes of morbidity and mortality, with rates that often surpass those of high-income countries^16^. In addition, many Latin American countries face a dual burden of disease, contending with both non-communicable diseases^17^ and ongoing infectious disease challenges^18^, including Zika, dengue, tuberculosis, and, more recently, COVID-19^19^. These health burdens are further compounded by inequities in access to care^20^, and disparities between urban and rural settings.

In this context, there is a critical need to make datasets from the region accessible to contribute to the global understanding of human care. To the best of our knowledge, few patient-level, longitudinal EHR datasets from primary care in Latin America have been made available to the research community. To address this gap, we introduce DiabetIA, a de-identified longitudinal EHR dataset derived from the primary care records of the Mexican Institute of Social Security (IMSS). The dataset includes 136,674 adult patients across six IMSS clinics in the state of Michoacán, capturing over 15 years of clinical care (2005–2020). DiabetIA provides structured data on demographics, diagnoses (ICD-10), prescriptions, laboratory results, and vital signs, summarized in two-year windows per patient.

An important feature of DiabetIA is that it captures longitudinal patterns of care rather than isolated clinical measurements of disease specific encounters. Its structure allows researchers to examine the coexistence and progression of chronic conditions, changes in anthropometric and physiological parameters, patterns of medication prescribing, and the use of primary care services over time. This may be particularly relevant for studying cardiometabolic multimorbidity in settings where access to laboratory testing and specialist care is variable and where routine clinical records represent an important source of real world evidence.

DiabetIA is designed to support a broad range of research applications, including the study of chronic disease progression, multimorbidity, prediction of T2D and its complications, and healthcare utilization in an underrepresented population. By making it freely available to the research community, we aim to advance more inclusive and equitable biomedical research and contribute to the international infrastructure for real-world evidence. The following sections describe the data source, cohort construction, variable extraction, standardization procedures, de-identification and technical validation of the DiabetIA resource.

## Methods

### Study Setting and Data Source

The dataset presented in this study was derived from the primary care electronic health records of the Instituto Mexicano del Seguro Social (IMSS), Mexico’s largest public healthcare provider. IMSS operates a nationwide, vertically integrated health system that covers more than 70 million beneficiaries, primarily through formal employment-based insurance. Its infrastructure includes primary care clinics (*Unidades de Medicina Familiar*, or UMFs), secondary hospitals, and tertiary referral centers. Primary care is the foundation of the IMSS care model, with UMFs serving as the first point of contact for the diagnosis, management, and follow-up of most chronic and acute conditions.

For this dataset, we extracted clinical records from six IMSS units with family medicine services located in the state of Michoacán, a western region of Mexico with a population of approximately 4.7 million inhabitants: four UMFs (No. 75, 76, 77 and 80) and two general hospitals with family medicine (HGZ/MF No. 2 and No. 12). The six participating units were selected in collaboration with institutional stakeholders based primarily on data availability and the feasibility of obtaining longitudinal electronic health records. They included a combination of medium and high volume family medicine services. Although these units provide information on routine primary care delivery within IMSS, their selection was not based on a probability sampling strategy and should not be interpreted as nationally representative of all IMSS facilities or beneficiaries.

The dataset includes longitudinal data from 2005 to 2020, encompassing over 15 years of routinely collected clinical information. We included all outpatient visits recorded in the SIMF database (*Sistema de Información en Medicina Familiar*), the primary care information system developed and used by IMSS. The source system records preventive care, chronic disease follow-up, new diagnoses, acute episodic care, child health and prenatal care; only adults (aged 18 years and older) without pregnancy-related conditions were included in this release (see Inclusion and Exclusion Criteria).

Each visit record includes structured data fields covering demographic characteristics, diagnostic codes, prescribed medications, laboratory results, vital signs, and visit metadata (e.g., timestamp, clinic ID). The dataset captures repeated encounters over time, allowing for the reconstruction of patient-level care trajectories in a real-world outpatient setting.

### Inclusion and Exclusion Criteria

The cohort was derived using a structured, multistep filtering process from the SIMF database (Figure 1). From an initial pool of records comprising 2,255,994 patients, duplicates were excluded using unique identifiers, yielding 1,729,070 distinct individuals. Patients were then filtered based on demographic completeness: 423,680 individuals were excluded due to invalid birthdates, being younger than 18 years on 31 December 2018, or missing sex data, resulting in 1,305,390 eligible adults.

**Figure 1.**
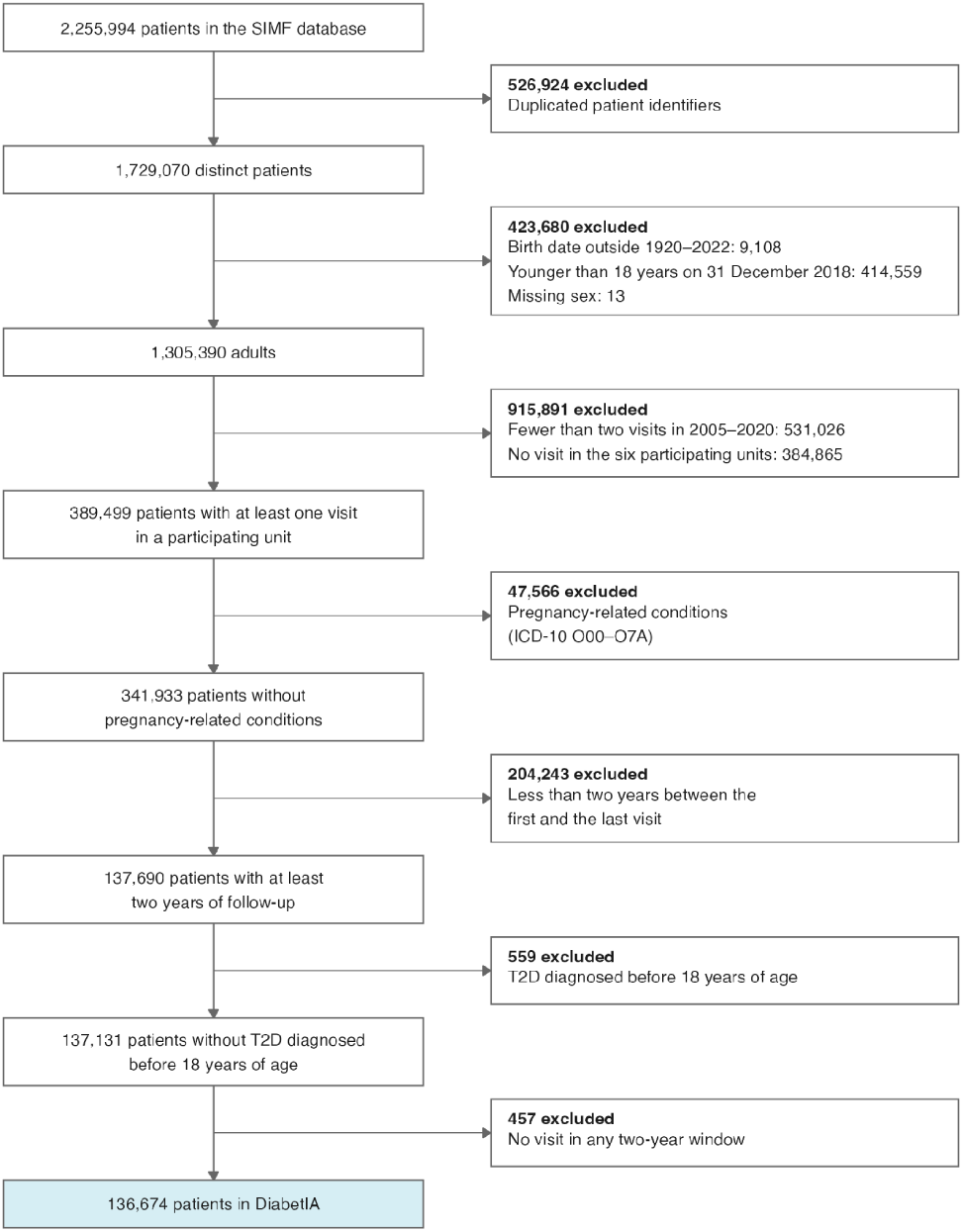
Selection of the DiabetIA cohort from the SIMF database. Exclusions were applied in the order shown; the final cohort comprises 136,674 adult patients from six family medicine units of IMSS in Michoacán (2005–2020).

To ensure consistency in clinical exposure and location, we excluded 531,026 individuals with fewer than two consultations during the observation window (2005–2020), and 384,865 patients who did not receive care at one of the six selected IMSS clinics in Michoacán. This yielded 389,499 patients with at least one recorded visit in a participating unit.

Patients with any record of pregnancy-related conditions were excluded (n=47,566; ICD-10 codes O00–O7A), leaving 341,933 individuals. A further 204,243 patients were removed due to insufficient follow-up (less than two years between the first and the last visit, so that every patient has at least one complete window of two years of follow-up and one outcome year), yielding 137,690 longitudinally monitored patients. In the final cohort, the time between the first and the last visit had a median of 3.8 years (interquartile range 2.9–4.8; maximum 16.0 years). All windows start at 18 years of age or later; 1,397 patients (1.0%) had their first recorded visit before that age.

To ensure age-consistent diagnoses, 559 patients with type 2 diabetes mellitus diagnosed before the age of 18 were excluded. Finally, 457 patients without a visit in any of their two-year windows, and therefore without a care unit or any clinical measurement in the windowed data, were excluded. The final cohort comprises 136,674 adult patients with structured, longitudinal primary care data suitable for real-world evidence generation.

The longitudinal eligibility criterion was applied to ensure that patients had sufficient temporal coverage for the construction of at least one observation window. For each eligible patient, the available visit history was used to define successive two-year follow-up windows, with a corresponding one year outcome period. Windows were initiated at one year intervals, resulting in a one-year overlap between consecutive observation windows. The availability of a visit within a window was assessed separately from the temporal eligibility of the patient; consequently, some eligible windows contained no recorded visits. These windows were retained in the released dataset and are identified through the corresponding visit and care unit fields.

### Data Extraction and Harmonization

Structured data were extracted from the electronic health record system using SQL-based queries and exported in tabular format for curation and standardization. Extracted variables were selected based on their relevance to longitudinal primary care analyses and their availability across the observation window.

The source records cover six domains: demographics (sex and date of birth); diagnoses, recorded with ICD-10 codes at each outpatient visit; anthropometry and vital signs (weight, height, body mass index [BMI], systolic and diastolic blood pressure, body temperature, heart rate and respiratory rate); laboratory results (glucose, HbA1c, lipids, creatinine, urea, uric acid and other commonly ordered assays), coded with LOINC and reported in UCUM units; prescribed drugs; and visit metadata (visit date, care unit and a de-identified patient ID). In the tabular release, these encounter-level records were aggregated into two-year windows per patient (see Data Records).

Data were extracted retrospectively from the SIMF database in three stages, following a medallion (bronze, silver and gold) architecture (Figure 2). First, IMSS engineers removed the tables with personal health information (PHI) before transferring the database to the research team (bronze); PHI embedded in the clinical narratives was removed with a pattern-matching algorithm, and the narratives are not part of this release. Second, the eligibility criteria were applied and the tables were aligned and harmonized (silver). Third, the harmonized data were merged, organized in two-year windows and aggregated into the tabular release (gold).

**Figure 2.**
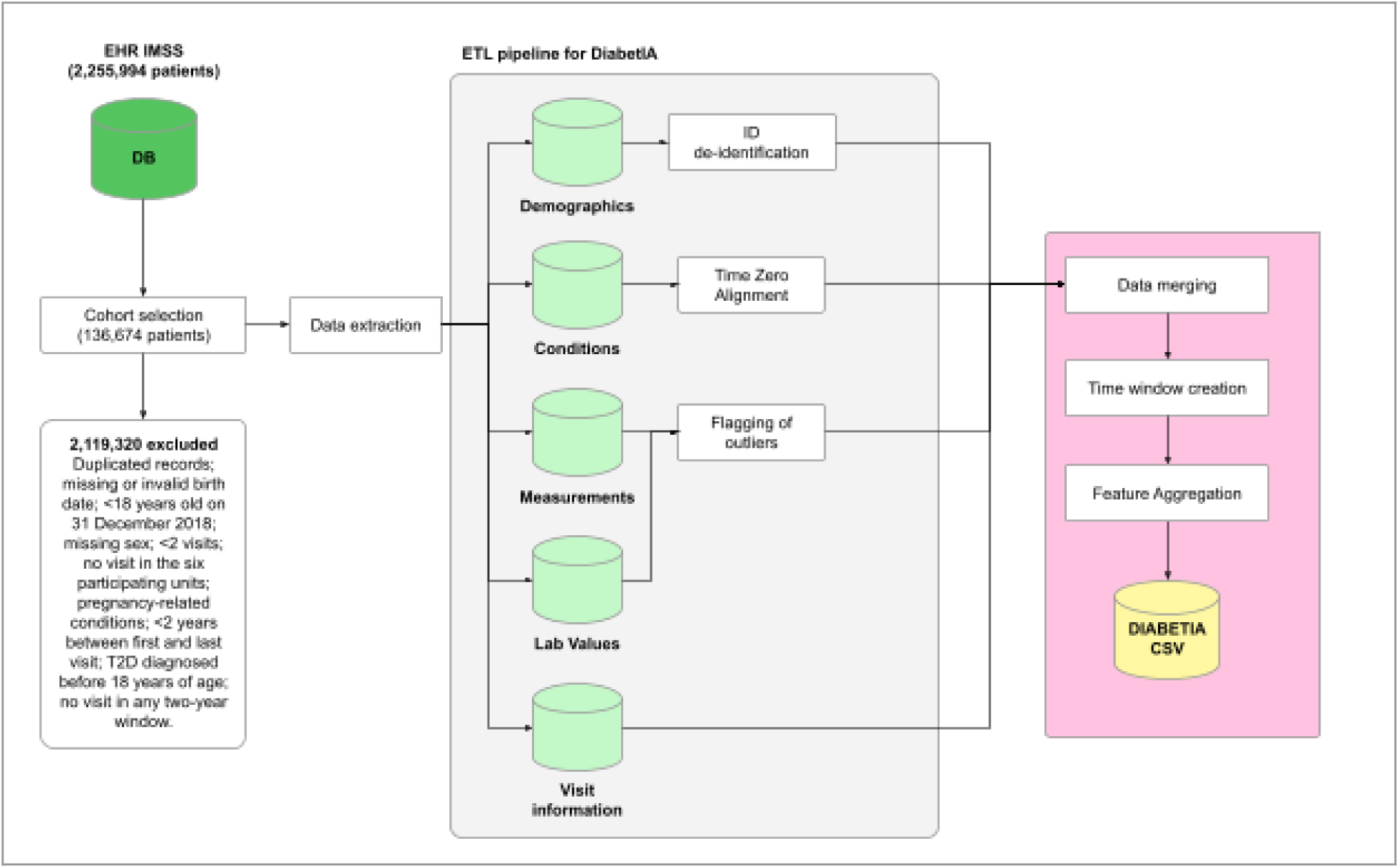
Overview of the development process for DiabetIA. Data is acquired from the IMSS electronic health record system (SIMF), and then processed in an ETL pipeline and aggregated into two-year windows in the tabular (CSV) release distributed to researchers.

### De-Identification and Privacy

All patient records in the DiabetIA dataset were de-identified prior to analysis and release, in accordance with Mexican data protection law and international standards for the use of health data in research. The de-identification process was designed to minimize the risk of re-identification while preserving the longitudinal structure necessary for analytical validity.

In Mexico, the protection of personal data held by public institutions is governed by the *Ley General de Protección de Datos Personales en Posesión de Sujetos Obligados* (LGPDPSO, enacted in March 2025). This law outlines the principles, rights, and responsibilities that public-sector entities must follow to ensure the lawful and secure handling of personal information. As a federal healthcare provider, the Mexican Institute of Social Security (IMSS) complies with these requirements through technical, administrative, and ethical safeguards, including data minimization, purpose limitation, access control, and de-identification. While the LGPDPSO does not specify de-identification standards equivalent to HIPAA’s Safe Harbor or Expert Determination methods, the approaches used in this project follow international best practices.

All direct personal identifiers, such as names, addresses, and national identification numbers, were removed from the dataset by the IMSS technical personnel. Patient records were de-identified, with author AGG serving as an honest broker. Dates of birth and clinical events were retained to preserve the temporal resolution needed for longitudinal analyses. No free-text fields or clinical notes are included in this version of DiabetIA. Outlying values were retained to preserve data fidelity and can be identified with the limits in Supplementary Table S4.

We assessed the residual risk of re-identification of the tabular release with k-anonymity, that is, the number of patients who share the same combination of quasi-identifiers. At the level of sex and year of birth, every combination was shared by at least 11 patients (median k = 996), the minimum cell size commonly used for the release of health data, and adding the care unit left only 8 patients (<0.01%) without a match (median k = 210). As expected for longitudinal data that keep exact dates, combinations that include the full date of birth are more distinctive: 8.6% of the patients were unique with sex and date of birth, and 54.2% and 99.4% when the care unit and the date of the first visit were added. This is the trade-off of retaining the temporal resolution needed for longitudinal analyses.

The re-identification risk assessment indicates that the preservation of exact dates and other quasi-identifiers may increase the uniqueness of individual records, particularly when multiple variables are considered jointly. Although direct identifiers were removed and patient identifiers were replaced with random codes, this process alone does not guarantee anonymity or eliminate the possibility of linkage attacks. Therefore, the results of the k-anonymity assessment should be interpreted as indicators of residual disclosure risk rather than as proof that individual re-identification is impossible.

The study protocol was reviewed and approved by the institutional review board of IMSS, through its Research and Ethics Committees (protocol R-2018-785-051). Because this is a retrospective study of secondary data, de-identified by author AGG as an honest broker, the committees waived informed consent, and IMSS approved the sharing of the dataset for research.

## Data Records

DiabetIA includes 136,674 patients who attended IMSS primary care units in Michoacán, a western Mexican state with 4.7 million inhabitants in 2020^21^. Table 1 describes the patients at their baseline window (the first two-year window with at least one visit), by T2D status and chronic complications; Supplementary Table S1 shows the same characteristics with diagnoses recorded in any window.

**Table 1.** Demographic, clinical and baseline biochemical characteristics of the study population.

|  | **Total** | **Without T2D** | **T2D without complications** | **T2D with complications** |
|---|---|---|---|---|
| **N** | 136,674 | 103,360 | 27,297 | 6,017 |
| **Sex** |  |  |  |  |
| Female | 79,098 (57.87%) | 59,444 (57.51%) | 16,304 (59.73%) | 3,350 (55.68%) |
| Male | 57,576 (42.13%) | 43,916 (42.49%) | 10,993 (40.27%) | 2,667 (44.32%) |
| **BMI (Weight kg/Height m2)** |  |  |  |  |
| Underweight | 1,257 (0.92%) | 1,182 (1.14%) | 54 (0.20%) | 21 (0.35%) |
| Normal weight | 32,431 (23.73%) | 27,425 (26.53%) | 3,824 (14.01%) | 1,182 (19.64%) |
| Overweight | 53,765 (39.34%) | 40,510 (39.19%) | 10,769 (39.45%) | 2,486 (41.32%) |
| Obesity (any class) | 44,132 (32.29%) | 29,324 (28.37%) | 12,482 (45.73%) | 2,326 (38.66%) |
| **Age at first window (years)** |  |  |  |  |
| 18 – 44 | 57,734 (42.24%) | 54,390 (52.62%) | 3,063 (11.22%) | 281 (4.67%) |
| 45 - 64 | 49,191 (35.99%) | 31,909 (30.87%) | 14,382 (52.69%) | 2,900 (48.20%) |
| 65 and older | 29,749 (21.77%) | 17,061 (16.51%) | 9,852 (36.09%) | 2,836 (47.13%) |
| **Comorbidities** |  |  |  |  |
| T2D (E11) | 33,314 (24.37%) | 0 (0.00%) | 27,297 (100.00%) | 6,017 (100.00%) |
| HTN (I10) | 50,638 (37.05%) | 25,594 (24.76%) | 20,093 (73.61%) | 4,951 (82.28%) |
| Dyslipidemia (E78) | 28,767 (21.05%) | 15,379 (14.88%) | 10,814 (39.62%) | 2,574 (42.78%) |
| Cardiovascular disease (I20–I25, I50) | 4,107 (3.00%) | 2,279 (2.20%) | 1,381 (5.06%) | 447 (7.43%) |
| Stroke (I60–I64, I69) | 1,198 (0.88%) | 648 (0.63%) | 368 (1.35%) | 182 (3.02%) |
| Any neoplasm (C00–D49) | 7,821 (5.72%) | 5,766 (5.58%) | 1,764 (6.46%) | 291 (4.84%) |
| **Chronic complications** |  |  |  |  |
| Diabetic nephropathy (E11.2) | 1,914 (1.40%) | 0 (0.00%) | 0 (0.00%) | 1,914 (31.81%) |
| Diabetic retinopathy (E11.3) | 1,829 (1.34%) | 0 (0.00%) | 0 (0.00%) | 1,829 (30.40%) |
| Diabetic neuropathy (E11.4) | 786 (0.58%) | 0 (0.00%) | 0 (0.00%) | 786 (13.06%) |
| Peripheral circulatory comp. (E11.5) | 2,537 (1.86%) | 0 (0.00%) | 0 (0.00%) | 2,537 (42.16%) |
| **Vital signs** |  |  |  |  |
| Systolic blood pressure (mmHg) | 117.6 (8.2); n=127,564 | 116.6 (8.2); n=94,547 | 120.2 (7.3); n=27,006 | 121.0 (7.4); n=6,011 |
| Diastolic blood pressure (mmHg) | 75.3 (5.8); n=127,571 | 74.9 (5.9); n=94,552 | 76.5 (5.3); n=27,008 | 76.7 (5.1); n=6,011 |
| **Biochemical** |  |  |  |  |
| Fasting glucose (mg/dL) | 122 (109–140); n=17,668 | – | 122 (110–140); n=13,944 | 122 (108–140); n=3,724 |
| Blood glucose (mg/dL) | 100 (90–120); n=49,460 | 94 (85–100); n=28,473 | 120 (104–147); n=16,698 | 126 (108–156); n=4,289 |
| HbA1c (%) | 6.8 (6.2–8.2); n=4,495 | – | 6.8 (6.1–8.1); n=3,662 | 7.1 (6.3–8.6); n=833 |
| Triglycerides (mg/dL) | 174 (131–234); n=11,144 | – | 175 (132–235); n=8,708 | 172 (130–233); n=2,436 |
| Total cholesterol (mg/dL) | 181 (157–206); n=18,132 | 184 (160–210); n=5,731 | 180 (156–204); n=9,910 | 178 (154–204); n=2,491 |
| Creatinine (mg/dL) | 0.80 (0.70–1.00); n=6,689 | 0.80 (0.70–1.00); n=5,259 | 0.80 (0.70–1.00); n=1,348 | 0.80 (0.70–1.40); n=82 |
| Urea (mg/dL) | 32.0 (25.8–39.8); n=2,244 | 31.7 (25.8–39.6); n=1,775 | 32.1 (25.6–39.9); n=426 | 35.8 (26.0–40.0); n=43 |
| Uric acid (mg/dL) | 5.5 (4.6–6.5); n=6,418 | 5.5 (4.6–6.5); n=5,041 | 5.5 (4.7–6.5); n=1,294 | 5.6 (4.6–7.0); n=83 |

T2D= Type 2 diabetes; HTN= Arterial hypertension; BMI= Body Mass Index. Underweight= BMI<18.5; Normal weight= BMI ≥18.5 and <25; Overweight= BMI ≥25 and <30; Obesity= BMI ≥30 (World Health Organization categories; see Supplementary Table S5). Age, BMI, vital signs, biochemical values and diagnoses were taken from the first two-year window with at least one visit of each patient (baseline window). Chronic diabetes-related complications were identified using the recorded ICD-10 codes E11.2, E11.3, E11.4 and E11.5. These codes were used as recorded in the source electronic health records and should be interpreted as diagnostic coding categories rather than as independently confirmed clinical outcomes. In particular, E11.5 was classified as diabetes with peripheral circulatory complications and was not assumed to represent diabetic foot in every case.

Women predominated in all groups (55.7–59.7%). T2D was recorded in 33,314 patients (24.4%), of whom 6,017 (18.1%) had chronic complications, mainly peripheral circulatory complications (42.2%), nephropathy (31.8%) and retinopathy (30.4%). Patients with T2D were older than those without T2D (90.0% versus 47.4% were 45 years or older). Overweight and obesity predominated in all groups (obesity 32.3% overall), and only 15.0% of the patients with T2D and 26.5% of those without T2D had a normal weight. Hypertension (37.1%) and dyslipidemia (21.0%) were the most frequent comorbidities, reaching 82.3% and 42.8% in patients with chronic complications. Among patients with T2D and a recorded value, the median fasting glucose was 122 mg/dL, HbA1c 6.8–7.1% and triglycerides 172–175 mg/dL.

Compared with other clinical data resources (Table 2), DiabetIA occupies a distinct niche. Population-wide primary care databases such as the Information System for Research in Primary Care (SIDIAP) in Catalonia, Spain^22^, and the Clinical Practice Research Datalink (CPRD) Aurum in England^23^ cover millions of patients, but they describe high-income European populations and are accessible only under institutional approval and, in the case of CPRD, a licence fee. MIMIC-IV^5^, one of the most widely used clinical databases, describes critical and emergency care in a single tertiary hospital in the United States and lacks longitudinal primary care information. Because MIMIC-IV is restricted to emergency department and intensive care admissions, its cohort represents acutely ill patients whose physiological measurements reflect acute decompensation rather than routine disease progression. In contrast, DiabetIA captures longitudinal clinical records from patients actively managing type 2 diabetes, avoiding acute-care spectrum bias and aligning directly with outpatient prevention of chronic complications. Within Mexico, the National Cancer Registry^24^ and the COVID-19 open data of the Ministry of Health^25^ are national in scope, but they focus on a single disease and do not follow patients over time; the cancer registry does not include follow-up care, laboratory data or outpatient medications, and the COVID-19 data have limited clinical detail outside acute respiratory episodes.

**Table 2.**
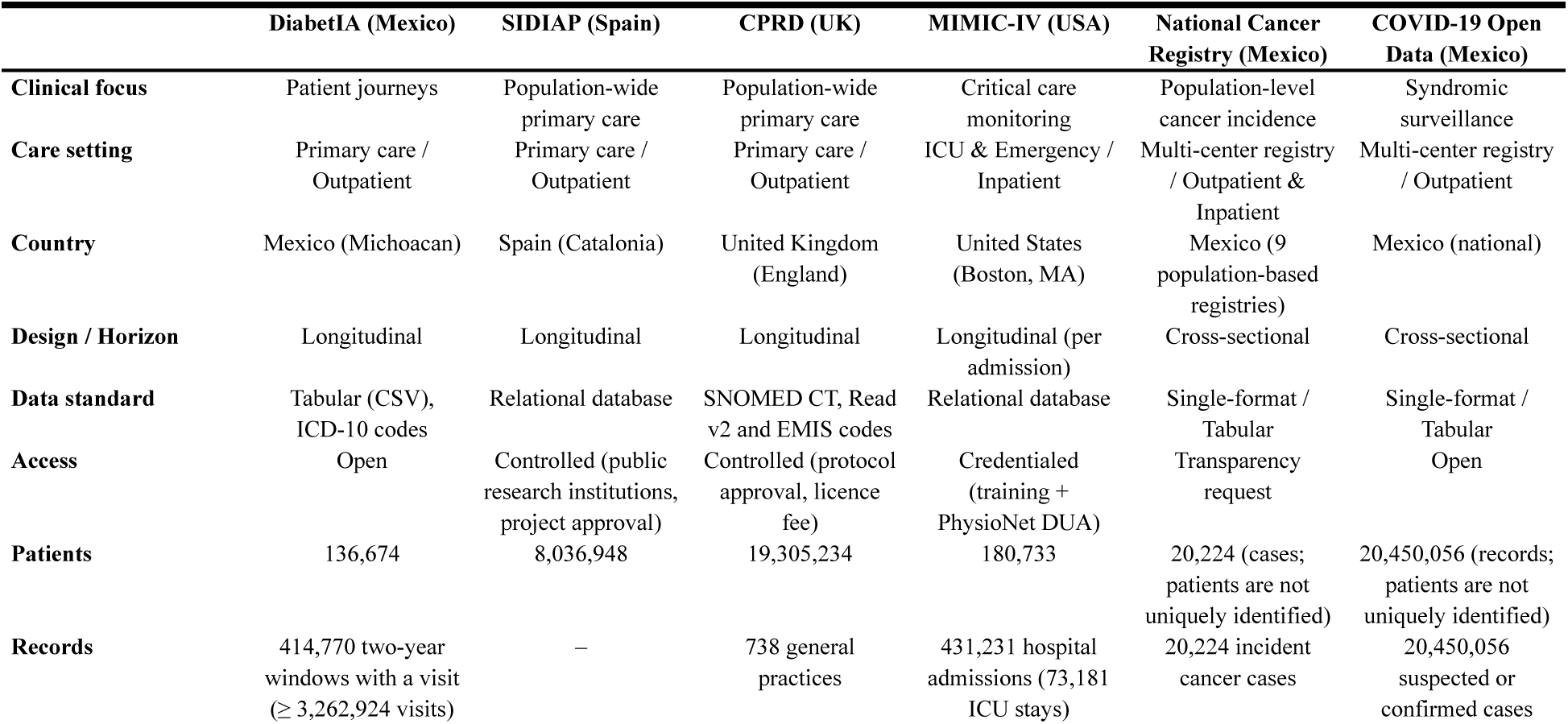
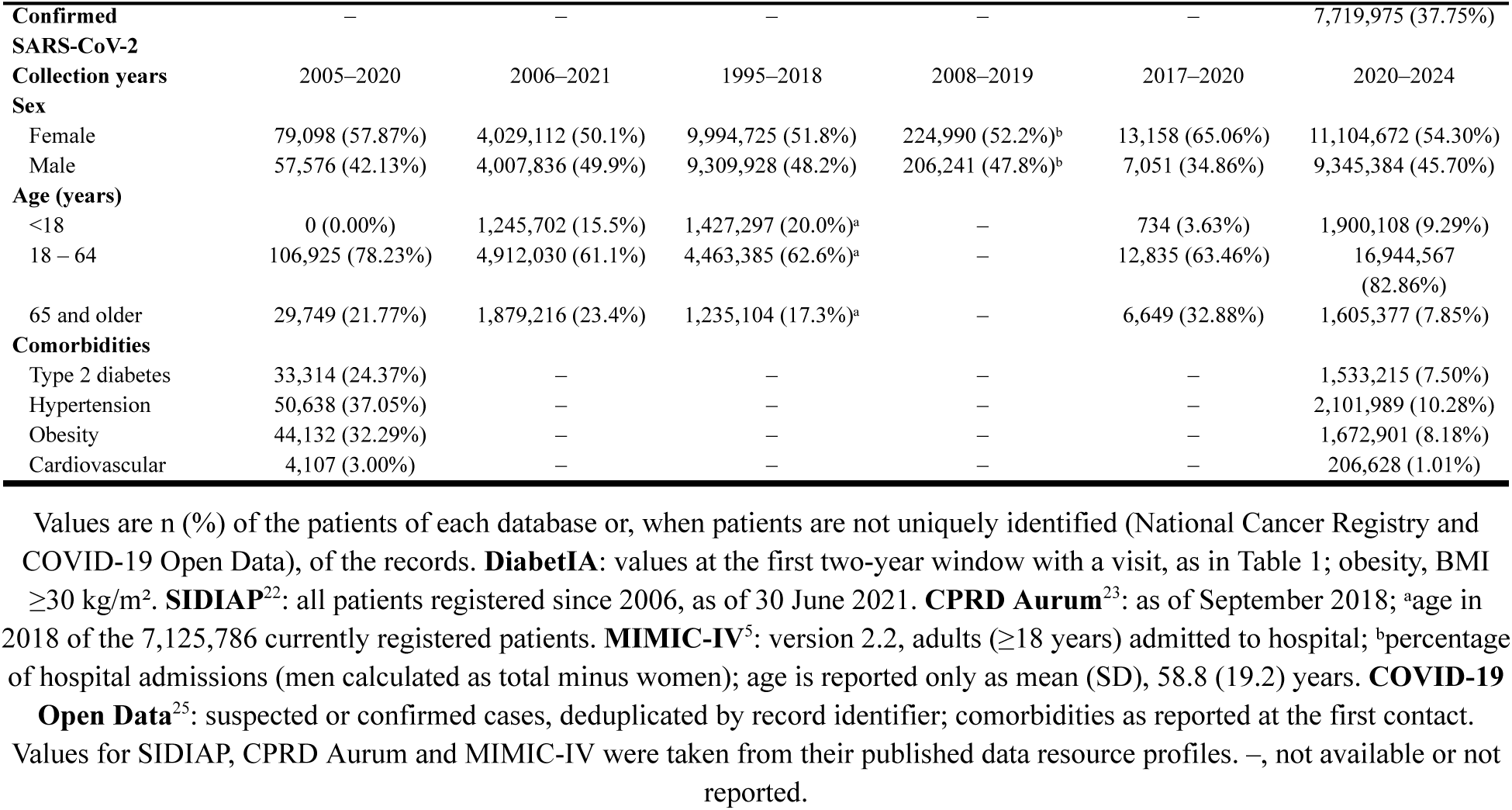
Comparison of DiabetIA with other clinical databases and registries.

|  | **DiabetIA (Mexico)** | **SIDIAP (Spain)** | **CPRD (UK)** | **MIMIC-IV (USA)** | **National Cancer Registry (Mexico)** | **COVID-19 Open Data (Mexico)** |
|---|---|---|---|---|---|---|
| **Clinical focus** | Patient journeys | Population-wide primary care | Population-wide primary care | Critical care monitoring | Population-level cancer incidence | Syndromic surveillance |
| **Care setting** | Primary care / Outpatient | Primary care / Outpatient | Primary care / Outpatient | ICU & Emergency / Inpatient | Multi-center registry / Outpatient & Inpatient | Multi-center registry / Outpatient |
| **Country** | Mexico (Michoacan) | Spain (Catalonia) | United Kingdom (England) | United States (Boston, MA) | Mexico (9 population-based registries) | Mexico (national) |
| **Design / Horizon** | Longitudinal | Longitudinal | Longitudinal | Longitudinal (per admission) | Cross-sectional | Cross-sectional |
| **Data standard** | Tabular (CSV), ICD-10 codes | Relational database | SNOMED CT, Read v2 and EMIS codes | Relational database | Single-format / Tabular | Single-format / Tabular |
| **Access** | Open | Controlled (public research institutions, project approval) | Controlled (protocol approval, licence fee) | Credentialed (training + PhysioNet DUA) | Transparency request | Open |
| **Patients** | 136,674 | 8,036,948 | 19,305,234 | 180,733 | 20,224 (cases; patients are not uniquely identified) | 20,450,056 (records; patients are not uniquely identified) |
| **Records** | 414,770 two-year windows with a visit (≥ 3,262,924 visits) | – | 738 general practices | 431,231 hospital admissions (73,181 ICU stays) | 20,224 incident cancer cases | 20,450,056 suspected or confirmed cases |
| **Confirmed SARS-CoV-2** | – | – | – | – | – | 7,719,975 (37.75%) |
| **Collection years** | 2005–2020 | 2006–2021 | 1995–2018 | 2008–2019 | 2017–2020 | 2020–2024 |
| **Sex** |  |  |  |  |  |  |
| Female | 79,098 (57.87%) | 4,029,112 (50.1%) | 9,994,725 (51.8%) | 224,990 (52.2%)<sup>b</sup> | 13,158 (65.06%) | 11,104,672 (54.30%) |
| Male | 57,576 (42.13%) | 4,007,836 (49.9%) | 9,309,928 (48.2%) | 206,241 (47.8%)<sup>b</sup> | 7,051 (34.86%) | 9,345,384 (45.70%) |
| **Age (years)** |  |  |  |  |  |  |
| <18 | 0 (0.00%) | 1,245,702 (15.5%) | 1,427,297 (20.0%)<sup>a</sup> | – | 734 (3.63%) | 1,900,108 (9.29%) |
| 18 – 64 | 106,925 (78.23%) | 4,912,030 (61.1%) | 4,463,385 (62.6%)<sup>a</sup> | – | 12,835 (63.46%) | 16,944,567 (82.86%) |
| 65 and older | 29,749 (21.77%) | 1,879,216 (23.4%) | 1,235,104 (17.3%)<sup>a</sup> | – | 6,649 (32.88%) | 1,605,377 (7.85%) |
| **Comorbidities** |  |  |  |  |  |  |
| Type 2 diabetes | 33,314 (24.37%) | – | – | – | – | 1,533,215 (7.50%) |
| Hypertension | 50,638 (37.05%) | – | – | – | – | 2,101,989 (10.28%) |
| Obesity | 44,132 (32.29%) | – | – | – | – | 1,672,901 (8.18%) |
| Cardiovascular | 4,107 (3.00%) | – | – | – | – | 206,628 (1.01%) |

Values are n (%) of the patients of each database or, when patients are not uniquely identified (National Cancer Registry and COVID-19 Open Data), of the records. **DiabetIA**: values at the first two-year window with a visit, as in Table 1; obesity, BMI ≥30 kg/m². **SIDIAP**<sup>22</sup>: all patients registered since 2006, as of 30 June 2021. **CPRD Aurum**<sup>23</sup>: as of September 2018; <sup>a</sup>age in 2018 of the 7,125,786 currently registered patients. **MIMIC-IV**<sup>5</sup>: version 2.2, adults (≥18 years) admitted to hospital; <sup>b</sup>percentage of hospital admissions (men calculated as total minus women); age is reported only as mean (SD), 58.8 (19.2) years. **COVID-19 Open Data**<sup>25</sup>: suspected or confirmed cases, deduplicated by record identifier; comorbidities as reported at the first contact. Values for SIDIAP, CPRD Aurum and MIMIC-IV were taken from their published data resource profiles. –, not available or not reported.

Among these resources, DiabetIA is the only one that is both longitudinal and openly available, and it describes a population with a high prevalence of T2D (24.4%), hypertension (37.1%) and obesity (32.3%), compared with 7.5%, 10.3% and 8.2% in the Mexican COVID-19 open data. DiabetIA therefore complements them with longitudinal outpatient data from an underrepresented Latin American population.

Figure 3 illustrates the longitudinal information available for each patient, using one patient selected automatically among those with long follow-up and complete anthropometry. Each column of the figure corresponds to one two-year window of the tabular release, so the trajectory is reconstructed directly from the rows of the patient. Diagnoses are cumulative and show the onset of T2D and of its chronic complications, whereas measurements are summarized in each window by their median and range, which shows both the long-term evolution of BMI, blood pressure and glucose and their variability between visits. The number of prescriptions of each drug group and the number of primary care visits show how treatment and the use of services change after the diagnosis of T2D and its complications. This type of trajectory can be used to study disease progression, the timing of complications and the response to treatment in routine care.

**Figure 3.**
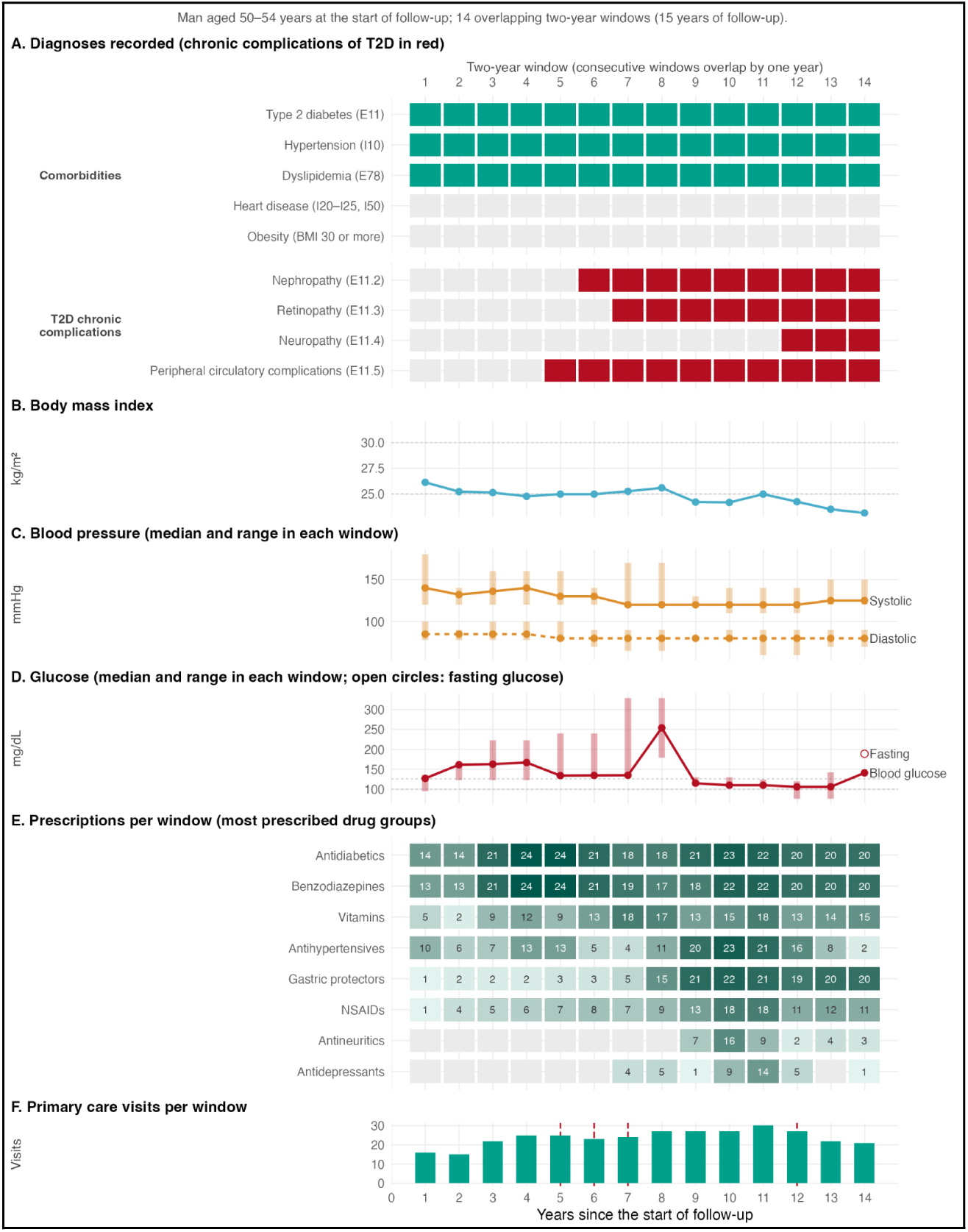
Longitudinal trajectory of one DiabetIA patient. Each column is a two-year window, placed at its centre in years since the start of follow-up; successive windows overlap by one year. (A) Diagnoses recorded up to each window, and obesity measured in the window; the chronic complications of T2D (nephropathy, retinopathy, neuropathy and peripheral circulatory complications) are shown in red; dashed vertical lines mark the window in which T2D (green, panels B–F, when it is diagnosed during follow-up) and each complication (red, panel F) were first recorded; (B) body mass index; (C) systolic and diastolic blood pressure, median and range of the values recorded in each window; (D) blood glucose (median and range) and fasting glucose; (E) number of prescriptions of the most prescribed drug groups; (F) number of primary care visits. The patient was selected automatically among those with long follow-up and complete anthropometry, prioritizing chronic complications that appeared during follow-up. To protect privacy, calendar dates are not shown and age is given in a 5-year band.

DiabetIA is owned by IMSS and is freely available for download, without registration, as a tabular release in CSV format from the National Informatics Ecosystem for Health (ENI Salud; see Data Availability). The tabular release is accompanied by a data dictionary (data_dictionary.xlsx) that describes the domain, data type and content of every column. Each row corresponds to an overlapping two-year window of one patient, identified by a de-identified patient identifier. Each window comprises a two-year follow-up period, in which the clinical information is summarized, and the following year (outcome period), in which the presence of T2D diagnoses is recorded as prediction targets. Each patient contributes between 1 and 15 successive windows (median 2, interquartile range 1–3; 38,355 patients, 28.1%, contribute a single window) that start one year apart, so consecutive windows overlap by one year (Supplementary Figure S1), for a total of 475,924 windows from the 136,674 patients, of which 414,770 included at least one visit. Follow-up periods span January 2005 to December 2020, and outcome periods extend to December 2021.

The 520 columns are grouped into six domains (Supplementary Table S6): demographics and visit information (17 columns, including sex, date of birth, age at the start of the window, care unit and number of visits); medical diagnoses (167 columns, indicating the presence of ICD-10 diagnoses during the follow-up period, individually or grouped by chapter); measurements (49 columns: weight, height, systolic and diastolic blood pressure, body temperature, heart rate and respiratory rate); laboratory results (128 columns, including glucose, HbA1c, lipids, creatinine, urea and uric acid); prescribed drugs (148 columns); and prediction targets (11 columns, corresponding to the ICD-10 codes E11 and E11.0–E11.9 in the outcome year). Measurements and laboratory results are summarized within each window by their mean, median, minimum, maximum, number of recorded values, standard deviation and slope over time, and prescriptions by their sum, mean, count and slope. Diagnoses behave cumulatively: once T2D was recorded for a patient, it was recorded in all later windows with a visit (see Technical Validation).

### Technical Validation

We assessed the quality of DiabetIA following the harmonized data quality framework of Kahn et al.^26^, which considers the completeness, plausibility and consistency of the data, and we also describe its temporal coverage. All analyses were run on the tabular release, in which each patient contributes successive two-year windows, and the code is publicly available (See Code Availability). Of the 475,924 two-year windows of the 136,674 patients, 414,770 (87.1%) included at least one primary care visit. The remaining 61,154 windows had no visit and therefore no recorded measurements, so completeness was computed over the windows with a visit.

#### Completeness

Sex and age were complete for all patients. Anthropometric measurements and vital signs were the most complete clinical variables: weight, height and BMI were recorded in 96.9% of the windows with a visit and at least once in 97.4% of patients, blood pressure in 93.8% of windows (95.8% of patients), body temperature in 95.3%, and heart and respiratory rates in 87.2–87.7%. Laboratory values were considerably sparser: blood glucose was recorded in 40.9% of windows, total cholesterol in 11.3%, fasting glucose in 11.2%, triglycerides in 7.0%, creatinine and uric acid in 4.0%, urea in 1.9%, and HbA1c in 2.1% of windows (4.4% of patients) (Supplementary Table S3).

Completeness depended on the clinical status of the patient in each window. Windows with a T2D diagnosis had more complete anthropometry and blood pressure (98.6–99.8%) than windows without T2D (91.8–95.9%), and blood glucose was recorded in 84.7% of the windows with T2D and chronic complications, compared with 29.0% of the windows without T2D. Fasting glucose, capillary glucose, HbA1c and triglycerides were recorded only in windows with a T2D diagnosis, which indicates that these fields are captured as part of diabetes follow-up rather than as general laboratory results. These variables are therefore missing not at random and should not be used to identify undiagnosed diabetes or to compare patients with and without T2D. In contrast, creatinine, urea and uric acid were less frequently recorded in windows with T2D and chronic complications (0.7–1.1%) than in windows without T2D (2.0–4.2%).

This differential measurement process has important implications for secondary analyses. The availability of laboratory results is influenced by clinical indication, disease recognition, follow-up practices, calendar period and care unit. Consequently, analyses restricted to patients with available laboratory measurements may be affected by selection bias, and complete case analyses may not provide unbiased estimates of associations in the full cohort.

#### Plausibility

Plausibility was assessed at two levels. First, the distribution of the within-window median of each variable (one value per patient, from the first window with a recorded value) was examined against its clinical reference range and categories (Supplementary Table S5; Figure 4). Weight, height, BMI, total cholesterol, urea and uric acid showed unimodal, physiological distributions, centred on the overweight range for BMI, whereas glucose, HbA1c and triglycerides were right-skewed, as expected in a population with a high prevalence of T2D. Second, the minimum and maximum values recorded in each window were compared with the most extreme values documented in the literature (Supplementary Table S4), and the windows beyond them were counted (Supplementary Table S2). No value of height, BMI, body temperature, heart rate, triglycerides, total cholesterol, creatinine or urea exceeded these limits. Values beyond the documented extremes were found for fasting glucose (298 windows, 0.64% of the windows with a value), the other glucose measurements (0.07–0.37%), respiratory rate (688 windows, 0.19%, all below 8 breaths/min), systolic and diastolic blood pressure (183 and 113 windows, ≤0.05%), weight (179 windows, 0.04%, all below 20 kg), HbA1c (9 windows, 0.10%) and uric acid (17 windows, 0.10%). These values were flagged but not removed, so that users can apply their own cleaning criteria.

**Figure 4.**
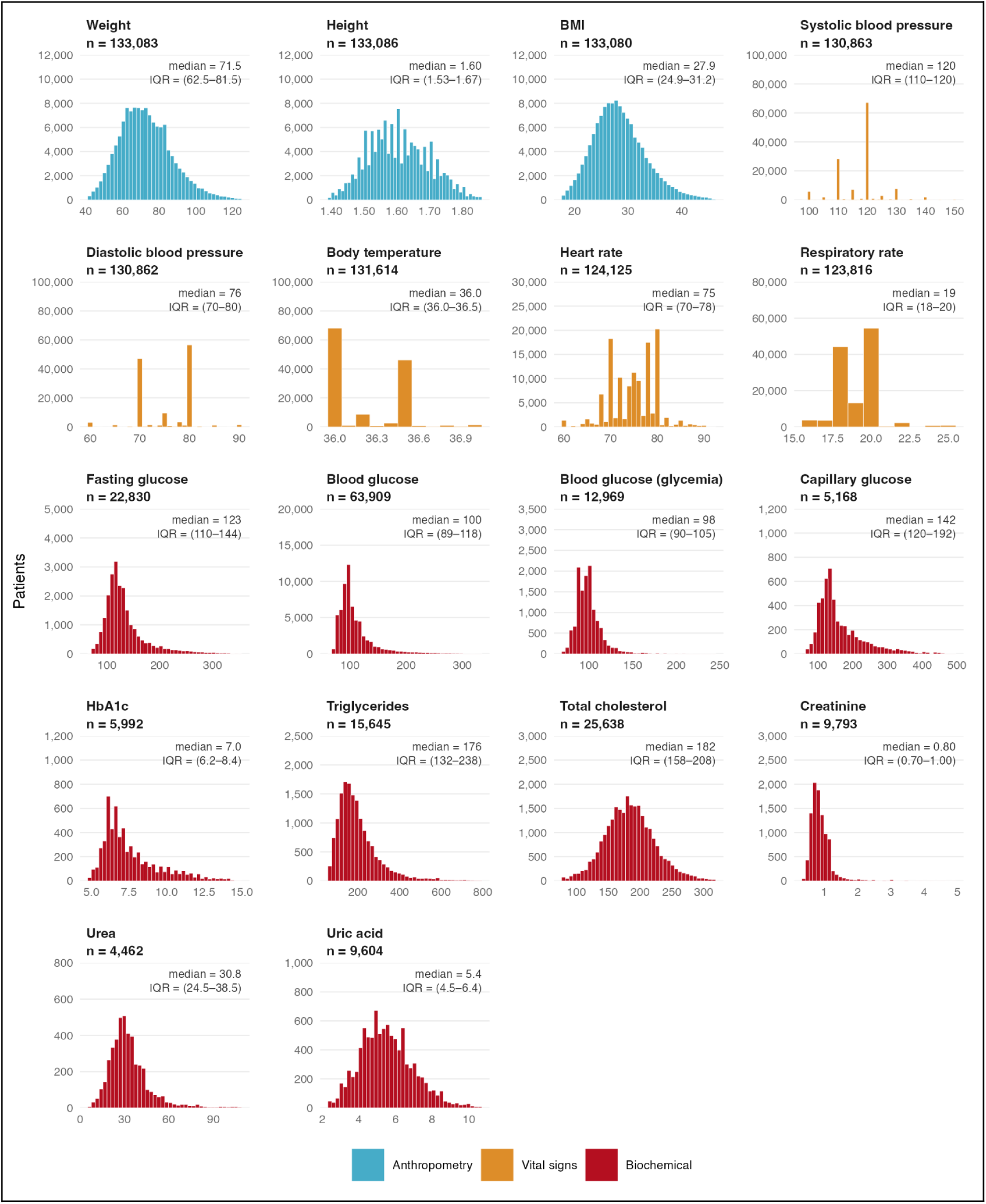
Distribution of anthropometric measurements, vital signs and laboratory values in DiabetIA. Each panel is a histogram of the number of patients, with one value per patient: the within-window median of the first two-year window with a recorded value of that variable. Because consecutive windows overlap by one year, counting windows would include some measurements twice and give more weight to patients with longer follow-up. n is the number of patients with a recorded value, and the text in each panel gives the median and interquartile range (IQR). For readability, the x-axis is limited to the 0.5th–99.5th percentiles of each variable; values outside this interval are summarized in Supplementary Table S2. Intermediate values in vital signs (e.g., 75 mmHg diastolic blood pressure) are medians of an even number of readings. Colours identify the group of variables. Units: weight, kg; height, m; BMI, kg/m²; blood pressure, mmHg; body temperature, °C; heart rate, beats/min; respiratory rate, breaths/min; HbA1c, %; all other biochemical values, mg/dL.

The distributions of the vital signs also revealed a strong terminal digit preference, a known feature of routinely collected primary care data: approximately 55% of the windows had a median systolic blood pressure of exactly 120 mmHg, 45% a diastolic blood pressure of 80 mmHg and 60% a body temperature of 36.0 °C, and respiratory rate concentrated at 18 and 20 breaths/min. Height clustered at multiples of 5 cm. Users should take this rounding into account when modelling small changes in these variables.

The observed concentration of measurements at commonly used reference values suggests substantial rounding and possible terminal digit preference during routine clinical recording. This pattern may reduce the ability to detect small physiological differences and may affect the estimation of short-term changes in blood pressure and body temperature. Accordingly, these variables are more suitable for analyses of broad clinical categories or larger differences than for detecting subtle longitudinal changes, unless measurement error is explicitly addressed.

#### Consistency

We evaluated 25 rules on dates, ages, diagnoses and the window structure (Supplementary Table S7). All windows had a two-year follow-up period followed by a one-year outcome period; the age recorded in each window agreed with the date of birth (within one year) and was between 18 and 110 years; a care unit was recorded if and only if the window had at least one visit; and every window with a visit fell within the recorded visit history of the patient. Successive windows of the same patient always started one year apart, so consecutive windows overlap by one year; the window numbering had gaps or did not start at 1 in 1,385 patients (1.0%). Among the 37,344 patients with a T2D diagnosis, the age at diagnosis agreed with the year of diagnosis and the date of birth, no diagnosis was made before 18 years of age, and once recorded, T2D was recorded again in all later windows with a visit. The year of diagnosis was later than the first window with a T2D record in 181 patients (0.5%). In 4,030 patients (10.8%), the year of diagnosis was recorded only from the window in which T2D first appeared; these are the patients diagnosed during follow-up (37,344 with T2D in any window versus 33,314 at their first window).

#### Temporal coverage

Follow-up periods spanned from January 2005 to December 2020, and outcome periods ended by December 2021 (Supplementary Table S8). Coverage was not uniform over time (Supplementary Figure S2): between 2005 and 2016, 17,000 to 21,000 windows with a visit started each year, compared with 97,625 in 2017 and 76,554 in 2018. This increase reflects the contribution of the care units: the two general hospitals with family medicine (HGZ/MF No. 2 and No. 12) contributed windows starting from 2005 to 2018, whereas UMF No. 75, 76 and 80 contributed windows starting from 2016 and UMF No. 77 from 2013, so their patients had a median of two windows, compared with four to five in the hospitals. Anthropometry and vital signs were recorded in 83% to 99% of the windows with a visit in every year, whereas laboratory results depended on the calendar period: HbA1c and capillary glucose were recorded only in windows starting in 2017 or later, and fasting glucose, triglycerides and total cholesterol were recorded in at most 9% of the windows starting before 2017 and in 11% to 24% of those starting in 2017 or 2018.

#### External comparison

We compared the prevalence of T2D, hypertension and obesity at the baseline window of the 131,939 patients aged 20 years or older with the 2018 National Health and Nutrition Survey (ENSANUT) in Michoacan^27^ (Supplementary Table S9). Obesity was similar in DiabetIA and in the survey (34.2% versus 31.6%, 95% confidence interval 26.5–37.3%; women 37.8% versus 37.4%, men 29.2% versus 24.4%). In contrast, recorded T2D (25.2% versus 9.9%) and hypertension (38.3% versus 19.2%) were two to two and a half times more frequent than the previous medical diagnoses reported in the survey, in both sexes and particularly from 40 years of age (T2D 28.8% versus 12.1% at 40–59 years and 42.7% versus 23.9% at 60 years or older). This difference is expected, because DiabetIA describes people who attend primary care, where chronic diseases are actively diagnosed and followed and diagnoses accumulate over the two-year baseline window, whereas the survey samples the general population and relies on self-reported diagnoses. The prevalence of both conditions increased with age as in the survey, and the agreement in measured obesity supports the validity of the anthropometric records.

This comparison should not be interpreted as a formal validation of population representativeness or as an assessment of individual level diagnostic agreement. Differences in sampling design, population composition, ascertainment methods and the accumulation of diagnoses over the two-year baseline window limit direct comparability. Similar obesity estimates provide evidence of epidemiological plausibility for this selected anthropometric measure, whereas the higher recorded prevalence of T2D and hypertension reflects, at least in part, the clinical selection and diagnostic characteristics of the primary care population.

## Usage Notes

DiabetIA can be downloaded directly from an institutional repository, which is further described in the Data Availability section. Users must not attempt to re-identify patients or to link the data with other sources. The tabular CSV file can be loaded with any statistical software, for example with pandas in Python or data.table in R. Users should read the data dictionary first, because each row is a two-year window and not a single visit: patient-level analyses should aggregate the windows of each patient (for example, using the first window with a visit as baseline), and window-level analyses should account for the repeated windows of each patient.

The prediction targets (E11 and E11.0–E11.9 in the outcome year) mostly identify prevalent rather than incident cases: about 95% of the windows with a positive target already had a T2D diagnosis during the follow-up period. Researchers building incidence prediction models should therefore restrict their analyses to windows without a previous T2D diagnosis. In addition, the E11 target is not the union of the E11.0–E11.9 targets, so each code should be used as recorded.

For predictive modelling, data partitioning should be performed at the patient level rather than at the window level. All windows belonging to the same patient should be assigned to the same training, validation or test partition to prevent information leakage arising from repeated and overlapping observations. When the objective is to predict incident T2D, only windows without a previous T2D diagnosis during the observation period should be considered eligible, and predictors should be restricted to information available before the outcome period. Temporal validation may also be considered to assess model performance across different calendar periods.

Laboratory results are “missing not at random” (MNAR) and should be analysed taking into account that most of them are recorded during the follow-up of T2D (Supplementary Table S3). Outlying values were not removed; users can apply the limits in Supplementary Table S4, which are also available in machine-readable form in the code repository (limits.csv), to exclude implausible values. Windows without a visit (12.9%) contain no measurements and can be excluded from most analyses. Analyses spanning the whole period should account for the care unit and the calendar year, because four of the six units contribute mainly to windows starting in 2016–2018 (Supplementary Table S8), and should take into account that successive windows of a patient overlap by one year.

The DiabetIA release is a tabular file in CSV format that can be analysed with open-source tools such as Python or R. The code used to generate the tables and figures of this article can serve as a starting point for loading and summarizing the tabular release (See Code Availability).

This release has several limitations. First, it includes six family medicine units in a single state and only IMSS beneficiaries, who are mostly formal-sector workers and their families; the cohort is therefore not representative of the uninsured population or of other regions of Mexico, and diagnosed T2D and hypertension were about twice as frequent as in the state population (Supplementary Table S9). Second, only adults were included, and patients with pregnancy-related conditions were excluded. Third, laboratory results are sparse and missing not at random: HbA1c was recorded in 2.1% of the windows with a visit, and only from 2017 onwards; their availability depends on the calendar period and the care unit; and fasting glucose, capillary glucose, HbA1c and triglycerides were recorded only in windows with a T2D diagnosis. Fourth, vital signs show a strong terminal digit preference, which limits the analysis of small changes in blood pressure or body temperature. Fifth, the dataset covers primary care only; hospital admissions and specialist care are not captured, and diagnoses recorded during routine care may be incomplete. Sixth, the last year of follow-up overlaps with the COVID-19 pandemic, which disrupted routine primary care in 2020. Finally, the clinical narratives written in the notes of each visit are not included in this release; their processing with natural language processing methods will be presented in a separate article.

Future releases will extend DiabetIA to more family medicine units and to other states served by IMSS, and will include patients under 18 years of age and pathologies beyond cardiometabolic diseases. In parallel, the dataset will be used to build predictive machine learning models, for example of T2D and its chronic complications, which will be reported in a separate article. We expect DiabetIA to support research on chronic disease trajectories, multimorbidity and health care use in an underrepresented population, and to serve as a reference for the release of other clinical datasets from Latin America.

## Data Availability

The DiabetIA dataset is available from the repository of the National Informatics Ecosystem for Health (ENI Salud) at https://repositorio-salud.conacyt.mx/jspui/handle/1000/296, where it can be downloaded without registration. The repository contains the tabular release in CSV format and its data dictionary.

## Code Availability

The code used to process the tabular release and to generate Tables and Figures is available at https://github.com/Pineda-Lab/diabetia_dataset under the MIT license, and archived in Zenodo^28^. It was written in Python (pandas 2.3.3, NumPy 2.0.2) and R (data.table, ggplot2). The repository contains only code; the data must be obtained as described in Data Availability.

## Acknowledgements

This project received partial funding from participating institutions and the National Council of Humanities, Sciences and Technologies (CONAHCyT, now the Secretariat of Science, Humanities, Technology and Innovation, SECIHTI) through its Institutional Fund for Regional Development for Scientific, Technological, and Innovation Development (FORDECyT) under the National Strategic Programs (ProNacEs), registered under number 10410.

## Author contributions

A.L.P. and C.A.A. conceived and designed the study. A.L.P. and K.M.F.M. supervised the data harmonization pipeline and drafted the initial manuscript. J.T. and D.S.Q. developed the software architecture of the extract, transform and load (ETL) process and executed the data extraction and harmonization. M.F.G., L.M.G.V. and K.M.F.M. contributed to the data validation, performed the statistical analyses, and curated the data quality reports. C.A.A. and A.G.G. acquired the funding, managed the regulatory and ethics approvals within the institution, provided clinical oversight, and critically reviewed the manuscript. All authors reviewed, edited, and approved the final version of the manuscript.

## Ethics statement

The study protocol was reviewed and approved by the Research and Ethics Committees of the Mexican Institute of Social Security (IMSS), under registration number R-2018-785-051. The study is a retrospective analysis of routinely collected electronic health records. All direct identifiers were removed by IMSS technical personnel, and the records were de-identified by author A.G.G., acting as an honest broker, before they were made available to the research team. Because the study used only secondary, de-identified data, the committees waived the requirement for informed consent. IMSS, as the owner of the data, approved the public release of the de-identified dataset for research purposes. The study was conducted in accordance with the Declaration of Helsinki and with the Mexican regulations on health research and on the protection of personal data held by public institutions (LGPDPSO).

