## Supplementary Material for "DiabetIA: A Longitudinal Electronic Health Record Dataset for Diabetes Research from Mexico"

**Supplementary Table S1. Demographic, clinical and baseline biochemical characteristics of the study population, with diagnoses present in any window of follow-up.**

| | Total | Without T2D | T2D without complications | T2D with complications |
|---|---|---|---|---|
| **N** | 136,674 | 99,330 | 27,411 | 9,933 |
| **Sex** | | | | |
| Female | 79,098 (57.87%) | 57,126 (57.51%) | 16,441 (59.98%) | 5,531 (55.68%) |
| Male | 57,576 (42.13%) | 42,204 (42.49%) | 10,970 (40.02%) | 4,402 (44.32%) |
| **BMI (Weight kg/Height m2)** | | | | |
| Underweight | 1,257 (0.92%) | 1,174 (1.18%) | 59 (0.22%) | 24 (0.24%) |
| Normal weight | 32,431 (23.73%) | 27,056 (27.24%) | 3,692 (13.47%) | 1,683 (16.94%) |
| Overweight | 53,765 (39.34%) | 39,063 (39.33%) | 10,689 (39.00%) | 4,013 (40.40%) |
| Obesity (any class) | 44,132 (32.29%) | 27,275 (27.46%) | 12,702 (46.34%) | 4,155 (41.83%) |
| **Age at first window (years)** | | | | |
| 18 – 44 | 57,734 (42.24%) | 53,076 (53.43%) | 3,803 (13.87%) | 855 (8.61%) |
| 45 - 64 | 49,191 (35.99%) | 30,044 (30.25%) | 14,091 (51.41%) | 5,056 (50.90%) |
| 65 and older | 29,749 (21.77%) | 16,210 (16.32%) | 9,517 (34.72%) | 4,022 (40.49%) |
| **Comorbidities** | | | | |
| T2D (E11) | 37,344 (27.32%) | 0 (0.00%) | 27,411 (100.00%) | 9,933 (100.00%) |
| HTN (I10) | 59,493 (43.53%) | 28,392 (28.58%) | 22,112 (80.67%) | 8,989 (90.50%) |
| Dyslipidemia (E78) | 40,585 (29.69%) | 21,240 (21.38%) | 13,570 (49.51%) | 5,775 (58.14%) |
| Heart or coronary disease (I20–I25, I50) | 6,390 (4.68%) | 3,177 (3.20%) | 1,982 (7.23%) | 1,231 (12.39%) |
| Stroke or cerebrovascular disease (I60–I64, I69) | 1,917 (1.40%) | 909 (0.92%) | 522 (1.90%) | 486 (4.89%) |
| Any neoplasm (C00–D49) | 12,507 (9.15%) | 8,684 (8.74%) | 2,767 (10.09%) | 1,056 (10.63%) |
| **Chronic complications** | | | | |
| Diabetic nephropathy (E11.2) | 3,631 (2.66%) | 0 (0.00%) | 0 (0.00%) | 3,631 (36.55%) |
| Diabetic retinopathy (E11.3) | 3,311 (2.42%) | 0 (0.00%) | 0 (0.00%) | 3,311 (33.33%) |
| Diabetic neuropathy (E11.4) | 1,977 (1.45%) | 0 (0.00%) | 0 (0.00%) | 1,977 (19.90%) |
| Peripheral circulatory complicationst (E11.5) | 4,600 (3.37%) | 0 (0.00%) | 0 (0.00%) | 4,600 (46.31%) |
| **Vital signs** | | | | |
| Systolic blood pressure (mmHg) | 117.6 (8.2); n=127,564 | 116.4 (8.1); n=90,803 | 119.9 (7.2); n=26,946 | 121.8 (8.3); n=9,815 |
| Diastolic blood pressure (mmHg) | 75.3 (5.8); n=127,571 | 74.8 (5.9); n=90,808 | 76.5 (5.3); n=26,948 | 77.3 (5.4); n=9,815 |
| **Biochemical** | | | | |
| Fasting glucose (mg/dL) | 122 (109–140); n=17,668 | – | 122 (110–140); n=13,056 | 122 (108–140); n=4,612 |
| Blood glucose (mg/dL) | 100 (90–120); n=49,460 | 93 (85–100); n=26,995 | 118 (101–140); n=15,471 | 127 (108–159); n=6,994 |
| HbA1c (%) | 6.8 (6.2–8.2); n=4,495 | – | 6.8 (6.1–8.1); n=3,520 | 7.0 (6.3–8.6); n=975 |
| Triglycerides (mg/dL) | 174 (131–234); n=11,144 | – | 175 (132–234); n=8,170 | 173 (130–234); n=2,974 |
| Total cholesterol (mg/dL) | 181 (157–206); n=18,132 | 184 (160–210); n=5,543 | 180 (157–204); n=9,527 | 179 (155–205); n=3,062 |
| Creatinine (mg/dL) | 0.80 (0.70–1.00); n=6,689 | 0.80 (0.70–1.00); n=5,094 | 0.80 (0.70–1.00); n=1,475 | 0.90 (0.70–1.32); n=120 |

| | Total | Without T2D | T2D without complications | T2D with complications |
|---|---|---|---|---|
| Urea (mg/dL) | 32.0 (25.8–39.8); n=2,244 | 31.7 (25.8–39.5); n=1,712 | 32.0 (25.7–39.8); n=459 | 33.0 (26.0–40.0); n=73 |
| Uric acid (mg/dL) | 5.5 (4.6–6.5); n=6,418 | 5.5 (4.6–6.5); n=4,885 | 5.5 (4.7–6.5); n=1,413 | 5.8 (4.8–7.0); n=120 |

T2D= Type 2 diabetes; HTN= Arterial hypertension; BMI= Body Mass Index. Underweight= BMI<18.5; Normal weight= BMI ≥18.5 and <25; Overweight= BMI ≥25 and <30; Obesity= BMI ≥30. Age, BMI, vital signs and biochemical values were taken from the first two-year window; diagnoses were present in any window of follow-up. T2D with complications= any diagnosis of E11.2, E11.3, E11.4 or E11.5. Categorical variables are n (%) relative to the total of each column. Continuous variables are median (interquartile range) of the patients with a recorded value (n).

**Supplementary Table S2. Plausibility of measurements and laboratory values: values recorded in DiabetIA compared with reference ranges and with extreme values documented in the literature.**

| | Windows with value, n (%) | Recorded minimum | Recorded maximum | Reference range | Extreme documented values (low – high) | Windows outside extreme values, n (%) | Ref. |
|---|---|---|---|---|---|---|---|
| **Anthropometry** | | | | | | | |
| Weight (kg) | 401,926 (96.9%) | 16 | 199.5 | – | 20 – 635 | 179 (0.04%) | 2, 10 |
| Height (m) | 401,946 (96.9%) | 1.02 | 2.19 | – | 0.546 – 2.72 | 0 (0.00%) | 11, 12 |
| BMI (kg/m²) | 401,907 (96.9%) | 8.44 | 86.95 | 18.5–24.9 | 6.7 – 100 | 0 (0.00%) | 2 |
| **Vital signs** | | | | | | | |
| Systolic blood pressure (mmHg) | 389,079 (93.8%) | 30 | 250 | 80–120 | 50 – 250 | 183 (0.05%) | 1 |
| Diastolic blood pressure (mmHg) | 389,113 (93.8%) | 30 | 199 | 60–80 | 20 – 150 | 113 (0.03%) | 1 |
| Body temperature (°C) | 395,210 (95.3%) | 25 | 41 | 36.5–37.5 | 13.7 – 46.5 | 0 (0.00%) | 13, 14 |
| Heart rate (beats/min) | 363,654 (87.7%) | 20 | 300 | 60–100 | 20 – 600 | 0 (0.00%) | 15, 16 |
| Respiratory rate (breaths/min) | 361,578 (87.2%) | 2 | 40 | 12–20 | 8 – 40+ | 688 (0.19%) | 17 |
| **Biochemical** | | | | | | | |
| Fasting glucose (mg/dL) | 46,403 (11.2%) | 10 | 2,732 | 70–100 | 10 – 1,000 | 298 (0.64%) | 3 |
| Blood glucose (mg/dL) | 169,439 (40.9%) | 2 | 999 | 70–100 (fasting) | 10 – 1,000 | 113 (0.07%) | 3 |
| Blood glucose (glycemia) (mg/dL) | 22,767 (5.5%) | 1.01 | 2,000 | 70–100 (fasting) | 10 – 1,000 | 69 (0.30%) | 3 |
| Capillary glucose (mg/dL) | 7,226 (1.7%) | 10 | 2,710 | 70–100 (fasting) | 10 – 1,000 | 27 (0.37%) | 3 |
| HbA1c (%) | 8,773 (2.1%) | 1.03 | 25 | <5.7 | 3 – 61.5 | 9 (0.10%) | 6 |
| Triglycerides (mg/dL) | 28,840 (7.0%) | 1.01 | 1,494 | <150 | > 0 – 10,000 | 0 (0.00%) | 9 |
| Total cholesterol (mg/dL) | 46,719 (11.3%) | 1.02 | 500 | <200 | > 0 – 1,000 | 0 (0.00%) | 3 |
| Creatinine (mg/dL) | 16,412 (4.0%) | 0.04 | 10 | 0.6–1.3 | > 0 – 20 | 0 (0.00%) | 3 |
| Urea (mg/dL) | 8,021 (1.9%) | 1.02 | 150 | 15–43 | > 0 – 642 | 0 (0.00%) | 3 |
| Uric acid (mg/dL) | 16,426 (4.0%) | 1.1 | 30 | 3.0–7.0 | > 0 – 20 | 17 (0.10%) | 3 |

Windows with value: % of the two-year windows with at least one visit. Recorded minimum and maximum are the lowest and highest values in any two-year window (BMI from the within-window median weight and height). Windows outside extreme values: windows whose minimum is below the lower limit or whose maximum is above the upper limit; values were flagged, not removed. "> 0": no extreme low value is documented, so only values ≤ 0 are flagged. Urea limits were converted from BUN (urea = BUN × 2.14). References: see the list below Supplementary Table S4.

**Supplementary Table S3. Completeness of demographic, anthropometric, vital sign and laboratory variables, by two-year window and type 2 diabetes status, and by patient.**

| | All windows (N=414,770) | Without T2D (N=298,967) | T2D without complications (N=82,467) | T2D with complications (N=33,336) | Patients, any window (N=136,674) |
|---|---|---|---|---|---|
| **Demographics** | | | | | |
| Sex | 414,770 (100.0%) | 298,967 (100.0%) | 82,467 (100.0%) | 33,336 (100.0%) | 136,674 (100.0%) |
| Age | 414,770 (100.0%) | 298,967 (100.0%) | 82,467 (100.0%) | 33,336 (100.0%) | 136,674 (100.0%) |
| **Anthropometry** | | | | | |
| Weight (kg) | 401,926 (96.9%) | 286,774 (95.9%) | 81,884 (99.3%) | 33,268 (99.8%) | 133,083 (97.4%) |
| Height (m) | 401,946 (96.9%) | 286,793 (95.9%) | 81,884 (99.3%) | 33,269 (99.8%) | 133,086 (97.4%) |
| BMI ($kg/m^2$) | 401,907 (96.9%) | 286,755 (95.9%) | 81,884 (99.3%) | 33,268 (99.8%) | 133,080 (97.4%) |
| **Vital signs** | | | | | |
| Systolic blood pressure (mmHg) | 389,079 (93.8%) | 274,524 (91.8%) | 81,345 (98.6%) | 33,210 (99.6%) | 130,863 (95.7%) |
| Diastolic blood pressure (mmHg) | 389,113 (93.8%) | 274,553 (91.8%) | 81,351 (98.6%) | 33,209 (99.6%) | 130,862 (95.7%) |
| Body temperature (°C) | 395,210 (95.3%) | 281,189 (94.1%) | 80,972 (98.2%) | 33,049 (99.1%) | 131,614 (96.3%) |
| Heart rate (beats/min) | 363,654 (87.7%) | 259,001 (86.6%) | 73,173 (88.7%) | 31,480 (94.4%) | 124,125 (90.8%) |
| Respiratory rate (breaths/min) | 361,578 (87.2%) | 257,183 (86.0%) | 72,951 (88.5%) | 31,444 (94.3%) | 123,816 (90.6%) |
| **Biochemical** | | | | | |
| Fasting glucose (mg/dL) | 46,403 (11.2%) | 0 (0.0%) | 31,905 (38.7%) | 14,498 (43.5%) | 22,830 (16.7%) |
| Blood glucose (mg/dL) | 169,439 (40.9%) | 86,601 (29.0%) | 54,587 (66.2%) | 28,251 (84.7%) | 63,909 (46.8%) |
| Blood glucose (glycemia) (mg/dL) | 22,767 (5.5%) | 16,334 (5.5%) | 5,622 (6.8%) | 811 (2.4%) | 12,969 (9.5%) |
| Capillary glucose (mg/dL) | 7,226 (1.7%) | 0 (0.0%) | 4,978 (6.0%) | 2,248 (6.7%) | 5,168 (3.8%) |
| HbA1c (%) | 8,773 (2.1%) | 0 (0.0%) | 6,986 (8.5%) | 1,787 (5.4%) | 5,992 (4.4%) |
| Triglycerides (mg/dL) | 28,840 (7.0%) | 0 (0.0%) | 19,866 (24.1%) | 8,974 (26.9%) | 15,645 (11.4%) |
| Total cholesterol (mg/dL) | 46,719 (11.3%) | 13,995 (4.7%) | 23,497 (28.5%) | 9,227 (27.7%) | 25,638 (18.8%) |
| Creatinine (mg/dL) | 16,412 (4.0%) | 12,472 (4.2%) | 3,649 (4.4%) | 291 (0.9%) | 9,793 (7.2%) |
| Urea (mg/dL) | 8,021 (1.9%) | 5,957 (2.0%) | 1,846 (2.2%) | 218 (0.7%) | 4,462 (3.3%) |
| Uric acid (mg/dL) | 16,426 (4.0%) | 12,314 (4.1%) | 3,758 (4.6%) | 354 (1.1%) | 9,604 (7.0%) |

T2D= Type 2 diabetes. Values are n (%) of windows (or patients) with a recorded value, relative to the N of each column. Window columns include only two-year windows with at least one visit; 61,154 of 475,924 windows (12.8%) had no visit and therefore no recorded measurements. A measurement is recorded in a window when it has at least one value during the two-year follow-up (within-window median not missing); BMI requires both weight and height. Window columns classify each window by the diagnoses recorded in that same window: T2D with complications= T2D and any diagnosis of E11.2, E11.3, E11.4 or E11.5. The last column counts patients with a recorded value in at least one window.

### Supplementary Table S4. Reference ranges and extreme values documented in the literature for the measurements and laboratory tests evaluated in the plausibility analysis.

| Test | Reference range | Extreme documented value, low | Extreme documented value, high | Ref. | Available in DiabetIA |
|---|---|---|---|---|---|
| **Anthropometry** | | | | | |
| Weight (kg) | – (see BMI) | 20 | 635 | 2, 10 | Yes |
| Height (m) | – | 0.546 | 2.72 | 11, 12 | Yes |
| BMI (kg/m²) | 18.5–24.9 | 6.7 | 100 | 2 | Yes |
| **Vital signs** | | | | | |
| Systolic blood pressure (mmHg) | 80–120 | 50 | 250 | 1 | Yes |
| Diastolic blood pressure (mmHg) | 60–80 | 20 | 150 | 1 | Yes |
| Body temperature (°C) | 36.5–37.5 | 13.7 | 46.5 | 13, 14 | Yes |
| Heart rate (beats/min) | 60–100 | 20 | 600 | 15, 16 | Yes |
| Respiratory rate (breaths/min) | 12–20 | 8 | 40+ | 17 | Yes |
| **Biochemical** | | | | | |
| Glucose (mg/dL) | 70–100 (fasting) | 10 | 1,000 (severe DKA/HHS) | 3 | Yes |
| HbA1c (%) | < 5.7 | 3.0 | 61.5 | 6 | Yes |
| Triglycerides (mg/dL) | < 150 | > 0 | 10,000 (severe hypertriglyceridemia) | 9 | Yes |
| Total cholesterol (mg/dL) | < 200 | > 0 | 1,000 | 3 | Yes |
| LDL cholesterol (mg/dL) | < 100 | < 20[a] | 190[a] | 7 | No |
| HDL cholesterol (mg/dL) | 40–80 | < 30[a] | 100[a] | 8 | No |
| Creatinine (mg/dL) | 0.6–1.3 | > 0 | 20 (end-stage renal disease) | 3 | Yes |
| Urea (mg/dL) | 15–43 (BUN 7–20 × 2.14) | > 0 | 642 (BUN 300 × 2.14) | 3 | Yes |
| Uric acid (mg/dL) | 3.0–7.0 | > 0 | 20 (severe tumor lysis syndrome) | 3 | Yes |
| Platelets (/µL) | 150,000–450,000 | 1,000 | 1,000,000 | 4 | No |
| eGFR (mL/min/1.73 m²) | 90–120 | 5 | 175 | 5 | No |

DKA= Diabetic ketoacidosis; HHS= Hyperglycemic hyperosmolar state; BUN= Blood urea nitrogen; eGFR= Estimated glomerular filtration rate. "> 0": no extreme low value is documented for this analyte, so only values ≤ 0 are considered implausible. Urea limits were converted from BUN (urea = BUN × 2.14). [a] Clinical risk thresholds, not extreme documented values; listed for reference only. Available in DiabetIA: tests recorded in the dataset; only these are evaluated in Supplementary Table S2.

9. Triglyceride Levels Greater Than 10,000 mg/dL in a 49-Year-Old Female without Evidence of Pancreatitis. Case Rep Endocrinol. 2019. PMCID: PMC6556268.

10. Guinness World Records. Heaviest man ever.

11. Guinness World Records. Tallest man ever.

12. Guinness World Records. Shortest man ever.

13. Gilbert M, Busund R, Skagseth A, Nilsen PA, Solbø JP. Resuscitation from accidental hypothermia of 13.7 °C with circulatory arrest. Lancet. 2000;355(9201):375-6.

14. Survival in a heat stroke victim with a core temperature in excess of 46.5 C. Ann Emerg Med. 1982.

15. Mouse Heart Rate in a Human: Diagnostic Mystery of an Extreme Tachyarrhythmia. Indian Pacing Electrophysiol J. 2012. PMCID: PMC3273956.

16. DiGiacinto J, Seladi-Schulman J. What is a normal vs. dangerous heart rate while resting and during activity? Healthline. 2024. https://www.healthline.com/health/dangerous-heart-rate

17. American Lung Association. Understanding vital signs: the importance of your respiratory rate. 2025. https://www.lung.org/blog/respiratory-rate-vital-signs

**Supplementary Table S5. Clinical categories and reference values for the measurements and laboratory tests recorded in DiabetIA.**

| Variable | Category | Values | Ref. |
|---|---|---|---|
| **Anthropometry** | | | |
| BMI (kg/m²) | Underweight | < 18.5 | 16 |
| | Normal weight | 18.5–24.9 | 16 |
| | Overweight | 25.0–29.9 | 16 |
| | Obesity class I | 30.0–34.9 | 16 |
| | Obesity class II | 35.0–39.9 | 16 |
| | Obesity class III | ≥ 40.0 | 16 |
| Weight (kg), height (m) | No clinical categories | Interpreted through BMI | – |
| **Vital signs** | | | |
| Blood pressure (mmHg) | Normal | SBP < 120 and DBP < 80 | 17 |
| | Elevated | SBP 120–129 and DBP < 80 | 17 |
| | Hypertension, stage 1 | SBP 130–139 or DBP 80–89 | 17 |
| | Hypertension, stage 2 | SBP ≥ 140 or DBP ≥ 90 | 17 |
| | Hypertensive crisis | SBP > 180 and/or DBP > 120 | 17 |
| | Hypertension (NOM-030 and ESH threshold) | SBP ≥ 140 or DBP ≥ 90 | 18 |
| Body temperature (°C) | Hypothermia | ≤ 35.0 | 19 |
| | Low | 35.1–36.0 | 19 |
| | Normal | 36.1–38.0 | 19 |
| | Fever | 38.1–39.0 | 19 |
| | High fever | ≥ 39.1 | 19 |
| Heart rate (beats/min) | Severe bradycardia | ≤ 40 | 19 |
| | Bradycardia | 41–50 | 19 |
| | Normal | 51–90 | 19 |
| | Mild tachycardia | 91–110 | 19 |
| | Tachycardia | 111–130 | 19 |
| | Severe tachycardia | ≥ 131 | 19 |
| Respiratory rate (breaths/min) | Severe bradypnoea | ≤ 8 | 19 |
| | Bradypnoea | 9–11 | 19 |
| | Normal | 12–20 | 19 |
| | Tachypnoea | 21–24 | 19 |
| | Severe tachypnoea | ≥ 25 | 19 |
| **Biochemical** | | | |
| Glucose (mg/dL)[a] | Hypoglycaemia, level 2 | < 54 | 21 |
| | Hypoglycaemia, level 1 | 54–69 | 21 |
| | Normal fasting glucose | 70–99 | 20 |
| | Prediabetes (impaired fasting glucose) | 100–125 (fasting) | 20 |
| | Diabetes | ≥ 126 (fasting) or ≥ 200 (random, with symptoms) | 20 |

| Variable | Category | Values | Ref. |
|---|---|---|---|
| HbA1c (%) | Normal | < 5.7 | 20 |
| | Prediabetes | 5.7–6.4 | 20 |
| | Diabetes | ≥ 6.5 | 20 |
| | Glycaemic goal for most adults with diabetes | < 7.0 | 21 |
| Total cholesterol (mg/dL) | Desirable | < 200 | 22 |
| | Borderline high | 200–239 | 22 |
| | High | ≥ 240 | 22 |
| Triglycerides (mg/dL) | Normal | < 150 | 22 |
| | Borderline high | 150–199 | 22 |
| | High | 200–499 | 22 |
| | Very high | ≥ 500 | 22 |
| Creatinine (mg/dL) | Reference interval (adults)[b] | 0.6–1.3 | 25 |
| | eGFR category G1 (normal or high) | eGFR ≥ 90 mL/min/1.73 $m^2$ | 23, 24 |
| | G2 (mildly decreased) | 60–89 | 23, 24 |
| | G3a (mildly to moderately decreased) | 45–59 | 23, 24 |
| | G3b (moderately to severely decreased) | 30–44 | 23, 24 |
| | G4 (severely decreased) | 15–29 | 23, 24 |
| | G5 (kidney failure) | < 15 | 23, 24 |
| Urea (mg/dL) | Reference interval (adults)[b] | 15–43 (BUN 7–20 × 2.14) | 25 |
| Uric acid (mg/dL) | Reference interval (adults)[b] | 3.0–7.0 | 25 |
| | Hyperuricaemia | > 6.8 (urate saturation) | 27 |
| | Treatment target in gout | < 6.0 | 26 |

BMI= Body mass index; SBP= Systolic blood pressure; DBP= Diastolic blood pressure; NOM-030= Mexican Official Standard NOM-030-SSA2-2009; ESH= European Society of Hypertension; eGFR= Estimated glomerular filtration rate; BUN= Blood urea nitrogen. Categories are listed to interpret the variables; except for the BMI categories (Table 1) and the reference lines of Figure 4 (BMI 25 and 30 kg/$m^2$; glucose 100 and 126 mg/dL), they were not applied to the data. Blood pressure categories follow the 2017 ACC/AHA guideline; the Mexican standard and the European guidelines define hypertension from 140/90 mmHg. The bands of temperature, heart rate and respiratory rate are those of the National Early Warning Score 2 (NEWS2), where the normal band scores 0; the category labels are descriptive. [a] Fasting thresholds apply to fasting glucose; blood, capillary and glycaemia values are not necessarily fasting, so only the random-glucose and hypoglycaemia thresholds apply to them. [b] Reference intervals depend on the analytical method and the laboratory. eGFR is not included in DiabetIA but can be estimated from creatinine, age and sex with the 2021 CKD-EPI equation.

19. Royal College of Physicians. National Early Warning Score (NEWS) 2: standardising the assessment of acute-illness severity in the NHS. Updated report of a working party. London: RCP; 2017.

20. American Diabetes Association Professional Practice Committee. 2. Diagnosis and classification of diabetes: Standards of Care in Diabetes—2025. Diabetes Care. 2025;48(Suppl 1):S27-S49.

21. American Diabetes Association Professional Practice Committee. 6. Glycemic goals and hypoglycemia: Standards of Care in Diabetes—2025. Diabetes Care. 2025;48(Suppl 1):S128-S145.

22. Expert Panel on Detection, Evaluation, and Treatment of High Blood Cholesterol in Adults. Executive summary of the Third Report of the National Cholesterol Education Program (NCEP) Expert Panel on Detection, Evaluation, and Treatment of High Blood Cholesterol in Adults (Adult Treatment Panel III). JAMA. 2001;285(19):2486-97.

23. Kidney Disease: Improving Global Outcomes (KDIGO) CKD Work Group. KDIGO 2024 clinical practice guideline for the evaluation and management of chronic kidney disease. Kidney Int. 2024;105(4S):S117-S314.

24. Inker LA, Eneanya ND, Coresh J, et al. New creatinine- and cystatin C-based equations to estimate GFR without race. N Engl J Med. 2021;385(19):1737-49.

25. Rifai N, Horvath AR, Wittwer CT, editors. Tietz textbook of clinical chemistry and molecular diagnostics. 6th ed. St. Louis: Elsevier; 2018.

26. FitzGerald JD, Dalbeth N, Mikuls T, et al. 2020 American College of Rheumatology guideline for the management of gout. Arthritis Care Res (Hoboken). 2020;72(6):744-60.

27. Dalbeth N, Gosling AL, Gaffo A, Abhishek A. Gout. Lancet. 2021;397(10287):1843-55.

### Supplementary Table S6. Data records: domains and columns of the DiabetIA tabular release.

| Domain | Columns, n | Content | Summary within each window | Coding |
|---|---|---|---|---|
| Demographics and visit information | 17 | Coded patient identifier, window number and dates, sex, age at the window, care unit, number of visits | One value per window | – |
| Medical diagnoses | 167 | Presence of ICD-10 diagnoses during the two-year follow-up period, individually or grouped by chapter | Presence | ICD-10 |
| Measurements | 49 | Weight, height, systolic and diastolic blood pressure, body temperature, heart rate, respiratory rate | Mean, median, minimum, maximum, count, SD, slope | UCUM units |
| Laboratory results | 128 | Glucose (fasting, capillary, blood), HbA1c, triglycerides, total cholesterol, creatinine, urea, uric acid and other commonly ordered tests | Mean, median, minimum, maximum, count, SD, slope | LOINC, UCUM units |
| Prescribed drugs | 148 | Prescriptions recorded during the follow-up period | Sum, mean, count, slope | IMSS catalog |
| Prediction targets | 11 | Diagnosis of T2D (E11) and its subcodes (E11.0–E11.9) in the year after the follow-up period | Presence | ICD-10 |
| **Total** | **520** | | | |

Each row of the tabular release is a two-year window of one patient (follow-up period) followed by a one-year outcome period. SD= standard deviation; slope= linear trend of the values over the window. The data dictionary (data_dictionary.xlsx) describes every column.

### Supplementary Table S7. Internal consistency of dates, ages, diagnoses and window structure.

| | Rule | Evaluated, n | Inconsistent, n (%) |
|---|---|---|---|
| **Window** | | | |
| Follow-up period order | x_start < x_end | 475,924 | 0 (0.00%) |
| Follow-up period length | x_end − x_start = 2 years (± 2 days) | 475,924 | 0 (0.00%) |
| Outcome period follows follow-up | y_start = x_end (± 1 day) | 475,924 | 0 (0.00%) |
| Outcome period length | y_end − y_start = 1 year (± 2 days) | 475,924 | 0 (0.00%) |
| Age vs date of birth | \|age_at_wx − age computed at x_start\| ≤ 1 year | 475,924 | 0 (0.00%) |
| Adult age | age_at_wx ≥ 18 years | 475,924 | 0 (0.00%) |
| Plausible age | age_at_wx ≤ 110 years | 475,924 | 0 (0.00%) |
| Visits and care unit | care unit recorded if and only if number of visits > 0 | 475,924 | 0 (0.00%) |
| Window within visit history | windows with a visit overlap first_cx – last_cx | 414,770 | 0 (0.00%) |
| Sex-specific diagnosis | no ovarian dysfunction (E28) in men | 210,415 | 0 (0.00%) |
| Sex-specific diagnosis | no testicular dysfunction (E29) in women | 265,509 | 0 (0.00%) |
| Persistence of T2D | T2D recorded again in later windows with a visit | 78,860 | 0 (0.00%) |
| **Patient** | | | |
| Window numbering | windows numbered 1, 2, …, k without gaps | 136,674 | 1,385 (1.01%) |
| Successive windows | x_start of successive windows 1 year apart (± 2 days) | 98,319 | 0 (0.00%) |
| Constant sex | same sex in all windows | 136,674 | 0 (0.00%) |
| Constant date of birth | same date of birth in all windows | 136,674 | 0 (0.00%) |
| Constant T2D diagnosis year and age | same dx_year_e11 and dx_age_e11 in all windows where they are recorded | 37,344 | 0 (0.00%) |
| T2D diagnosis year in all windows | dx_year_e11 recorded in every window once it is recorded in one | 37,344 | 4,030 (10.79%) |
| Visit history order | first_cx ≤ last_cx | 136,674 | 0 (0.00%) |
| Age at T2D diagnosis | \|dx_age_e11 − (dx_year_e11 − year of birth)\| ≤ 1 year | 37,344 | 0 (0.00%) |
| Adult-onset T2D | dx_age_e11 ≥ 18 years | 37,344 | 0 (0.00%) |

| | Rule | Evaluated, n | Inconsistent, n (%) |
|---|---|---|---|
| T2D diagnosis year within life and observation | year of birth ≤ dx_year_e11 ≤ year of last y_end | 37,344 | 0 (0.00%) |
| T2D diagnosis with year | patients with T2D recorded in a window have dx_year_e11 | 37,344 | 0 (0.00%) |
| T2D diagnosis year before first record | dx_year_e11 ≤ year of x_end of the first window with T2D | 37,344 | 181 (0.48%) |
| Diagnosis year with T2D record | patients with dx_year_e11 have T2D recorded in some window | 37,344 | 0 (0.00%) |

Each rule was evaluated on the windows (or patients) with the information required; inconsistent records were flagged, not removed. Follow-up period= x_start to x_end; outcome period= y_start to y_end. Ages were computed in completed years at the start of the window (x_start); differences of up to 1 year were accepted because of rounding conventions. T2D= type 2 diabetes (diabetes_mellitus_type_2 diagnosis column; dx_year_e11 and dx_age_e11 for the year and age at diagnosis). Persistence of T2D is descriptive: a chronic diagnosis is expected to be recorded again in later windows with a visit.

### Supplementary Table S8. Temporal coverage of DiabetIA by care unit.

| | Patients, n | Windows with a visit, n | Windows per patient | Years |
|---|---|---|---|---|
| **Care unit** | | | | |
| HGZ con M.F. No. 12 | 22,531 | 124,309 | 4 (2–9) | 2005–2018 |
| HGZ con M.F. No. 2 | 24,945 | 146,921 | 5 (2–9) | 2005–2018 |
| UMF No. 75 | 28,271 | 45,469 | 2 (1–2) | 2016–2018 |
| UMF No. 76 | 9,741 | 15,891 | 2 (1–2) | 2016–2018 |
| UMF No. 77 | 12,614 | 20,631 | 2 (1–2) | 2013–2018 |
| UMF No. 80 | 38,572 | 61,549 | 2 (1–2) | 2016–2018 |
| **Total** | 136,674 | 414,770 | 2 (1–2) | 2005–2018 |

Only two-year windows with at least one visit are counted. Windows per patient: median (interquartile range) of the windows with a visit of each patient in that care unit. Years: calendar years of the start of the first and last window (x_start). Successive windows of a patient start one year apart and overlap by one year. Follow-up periods span 2005-01-03 to 2020-12-30, and outcome periods end by 2021-12-30. Recorded visit history (first_cx – last_cx): 2005-01-03 to 2020-12-31.

**Supplementary Table S9. Prevalence of type 2 diabetes, hypertension and obesity in DiabetIA compared with the 2018 National Health and Nutrition Survey (ENSANUT) in Michoacán.**

| | DiabetIA, % (n) | ENSANUT 2018 Michoacán, % (95% CI) |
|---|---|---|
| **Type 2 diabetes** | | |
| 20–39 years | 3.8 (n=42,345) | 1.9 (1.0–3.6) |
| 40–59 years | 28.8 (n=46,957) | 12.1 (9.2–15.6) |
| 60 years and older | 42.7 (n=42,637) | 23.9 (19.0–29.7) |
| Total (20 years and older) | 25.2 (n=131,939) | 9.9 (8.5–11.6) |
| Women, Total (20 years and older) | 25.7 (n=76,456) | 11.8 (9.5–14.5) |
| Men, Total (20 years and older) | 24.6 (n=55,483) | 7.8 (6.1–9.9) |
| **Hypertension** | | |
| 20–39 years | 5.6 (n=42,345) | 8.0 (5.7–11.1) |
| 40–59 years | 39.9 (n=46,957) | 20.1 (16.2–24.5) |
| 60 years and older | 69.2 (n=42,637) | 42.1 (35.7–48.7) |
| Total (20 years and older) | 38.3 (n=131,939) | 19.2 (16.9–21.6) |
| Women, Total (20 years and older) | 40.3 (n=76,456) | 23.7 (20.5–27.3) |
| Men, Total (20 years and older) | 35.6 (n=55,483) | 13.7 (11.1–16.9) |
| **Obesity** | | |
| 20–39 years | 27.0 (n=39,721) | – |
| 40–59 years | 41.4 (n=45,575) | – |
| 60 years and older | 33.3 (n=41,976) | – |
| Total (20 years and older) | 34.2 (n=127,272) | 31.6 (26.5–37.3) |
| Women, Total (20 years and older) | 37.8 (n=74,549) | 37.4 (29.0–46.7) |
| Men, Total (20 years and older) | 29.2 (n=52,723) | 24.4 (19.2–30.5) |

DiabetIA: patients aged 20 years or older at their baseline window (first two-year window with a visit, as in Table 1); diabetes and hypertension are diagnoses recorded in that window (ICD-10 E11 and I10), and obesity is BMI ≥30 kg/m² measured in that window (among patients with BMI). ENSANUT 2018, Michoacán: adults aged 20 years or older; diabetes and hypertension by previous medical diagnosis reported in the survey, and obesity measured (BMI ≥30 kg/m²); percentage (95% confidence interval), from Tables 4.15, 4.16 and 5.21 of the state report (Instituto Nacional de Salud Pública, 2020). DiabetIA describes people who attend IMSS primary care, not the general population, and its baseline windows span 2005–2018. – = not reported by age group.

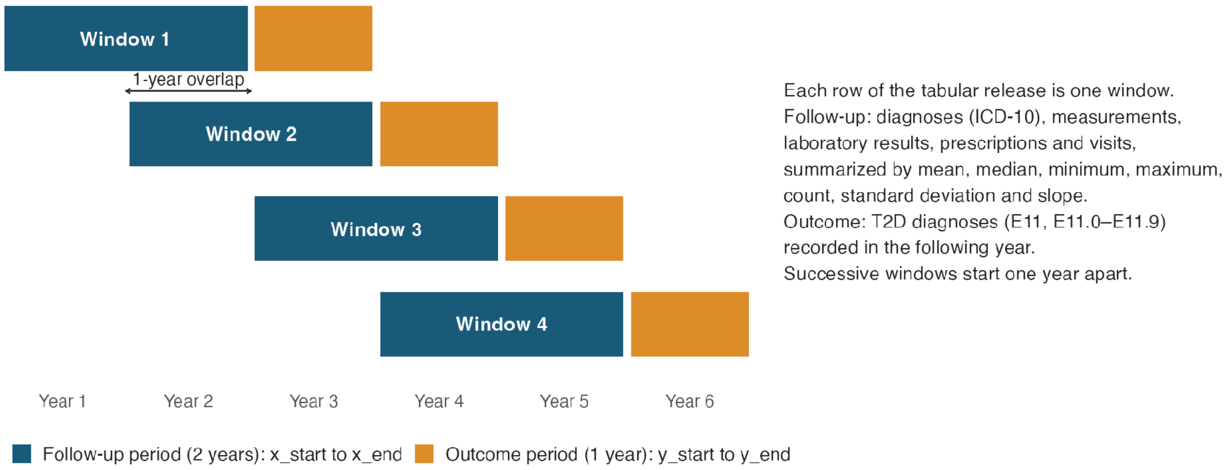


**Supplementary Figure S1.** Design of the DiabetIA windows. Each row of the tabular release is a two-year window of one patient: the follow-up period (x_start to x_end), in which diagnoses, measurements, laboratory results, prescriptions and visits are summarized, and the following year (outcome period, y_start to y_end), in which the T2D diagnoses (E11, E11.0–E11.9) used as prediction targets are recorded. Successive windows of a patient start one year apart, so consecutive windows overlap by one year. The schematic shows four windows and contains no patient data.

| Group | Variable | 2005 | 2006 | 2007 | 2008 | 2009 | 2010 | 2011 | 2012 | 2013 | 2014 | 2015 | 2016 | 2017 | 2018 |
|---|---|---|---|---|---|---|---|---|---|---|---|---|---|---|---|
| **Windows** | Windows with a visit (×1,000) | 17 | 18 | 19 | 19 | 20 | 21 | 21 | 21 | 21 | 21 | 21 | 21 | 98 | 77 |
| **Anthropometry** | Weight (kg) | 98 | 99 | 98 | 97 | 96 | 95 | 96 | 97 | 96 | 95 | 95 | 95 | 97 | 98 |
| | Height (m) | 98 | 99 | 98 | 98 | 96 | 95 | 96 | 97 | 96 | 95 | 95 | 95 | 97 | 98 |
| | BMI (kg/m²) | 98 | 99 | 98 | 97 | 96 | 95 | 96 | 97 | 96 | 95 | 95 | 95 | 97 | 98 |
| **Vital signs** | Systolic blood pressure (mmHg) | 93 | 93 | 93 | 91 | 90 | 90 | 92 | 93 | 92 | 92 | 92 | 93 | 96 | 97 |
| | Diastolic blood pressure (mmHg) | 93 | 93 | 93 | 91 | 90 | 90 | 92 | 93 | 92 | 92 | 92 | 93 | 96 | 97 |
| | Body temperature (°C) | 95 | 96 | 97 | 96 | 94 | 93 | 94 | 95 | 94 | 94 | 94 | 94 | 96 | 97 |
| | Heart rate (beats/min) | 88 | 88 | 86 | 85 | 85 | 84 | 88 | 90 | 89 | 89 | 90 | 90 | 88 | 87 |
| | Respiratory rate (breaths/min) | 85 | 86 | 86 | 84 | 84 | 83 | 88 | 89 | 89 | 89 | 89 | 90 | 88 | 87 |
| **Biochemical** | Fasting glucose (mg/dL) | 0 | 0 | 2 | 6 | 7 | 7 | 7 | 6 | 5 | 5 | 4 | 6 | 18 | 23 |
| | Blood glucose (mg/dL) | 40 | 37 | 40 | 43 | 41 | 37 | 38 | 39 | 40 | 40 | 44 | 47 | 41 | 42 |
| | Blood glucose (glycemia) (mg/dL) | 0 | 0 | 1 | 3 | 4 | 5 | 4 | 2 | 1 | 1 | 0 | 3 | 9 | 12 |
| | Capillary glucose (mg/dL) | 0 | 0 | 0 | 0 | 0 | 0 | 0 | 0 | 0 | 0 | 0 | 0 | 3 | 5 |
| | HbA1c (%) | 0 | 0 | 0 | 0 | 0 | 0 | 0 | 0 | 0 | 0 | 0 | 0 | 4 | 6 |
| | Triglycerides (mg/dL) | 0 | 0 | 1 | 4 | 5 | 5 | 5 | 3 | 2 | 2 | 1 | 3 | 11 | 15 |
| | Total cholesterol (mg/dL) | 0 | 0 | 2 | 6 | 8 | 9 | 8 | 5 | 3 | 3 | 2 | 6 | 19 | 24 |
| | Creatinine (mg/dL) | 0 | 0 | 0 | 1 | 2 | 3 | 2 | 1 | 1 | 0 | 0 | 2 | 7 | 9 |
| | Urea (mg/dL) | 0 | 0 | 0 | 1 | 2 | 2 | 2 | 1 | 1 | 0 | 0 | 2 | 3 | 4 |
| | Uric acid (mg/dL) | 0 | 0 | 0 | 2 | 3 | 3 | 3 | 1 | 1 | 1 | 0 | 2 | 7 | 9 |

Year of window start

Windows with a value (%): 0% 25% 50% 75% 100%

**Supplementary Figure S2.** Temporal coverage of DiabetIA. Percentage of the two-year windows with a visit that have a recorded value of each variable, by calendar year of the start of the window. The top row shows the number of windows with a visit that started each year (thousands). Successive windows of a patient start one year apart. Sex and age are recorded in every window and are not shown.
